# The children left behind – the cumulative impact of congenital anomalies, long-term conditions and poverty on educational attainment in Wales: a population databank linkage study

**DOI:** 10.64898/2026.04.01.26349936

**Authors:** Ieuan Scanlon, Anna Rawlings, David Tucker, Daniel S. Thayer, Hywel T. Evans, Joanne Given, Sam Jones, Maria Loane, Chloe Morgan, Joan K. Morris, Sue Jordan

## Abstract

**Background:** Education outcomes predict life chances. However, poverty, ill-health and disability are barriers to achievement. We examined determinants of academic attainment of children with and without major congenital anomalies in state-funded mainstream schools at ages 11 and 16 (key stages [KS] 2 and 4).

**Methods and Findings:** Routinely collected electronic records for children born in Wales 01/01/1998–31/12/2007 until 31/12/2019 were linked in the Secure Anonymised Information Linkage (SAIL) Databank. Education outcomes were explored using logistic regression, adjusting for: anomalies, maternal and child deprivation, prescribing, hospitalisation, gestation length, child’s sex, and special education needs (SEN) provision.

Children with anomalies were less likely to achieve academic standards: however, attainment was more closely associated with affluence. At age 11, 81.87% (7167/8754) with and 93.80% (232,450/247,814) without anomalies passed (odds ratio [OR] 0.30, 95% confidence intervals [CI] 0.28-0.32). At age 16, 46.76% (2070/4427) with and 56.10% (69,732/124,300) without anomalies achieved 5 General Certificates of Secondary Education (GCSEs) at grades C–A* including English/Welsh, Maths and Science (EWMS) (OR 0.69, 0.65–0.73). Discrepancies narrowed in adjusted analyses, particularly when SEN provision was accounted: aOR 0.72 (0.66–0.78) at KS2, and aOR 0.93, (0.87–1.00) for 5 GCSEs C–A* with EWMS. These GCSEs were achieved by 29.65% (307/1034) children with anomalies and 38.42% (10,875/28,305) of unaffected children in the most deprived quintile†: in the most affluent quintile, figures were 67.57% (547/810) and 74.98% (16,978/22,644). Children with anomalies, receiving maximum SEN support, eligible for Free School Meals (FSM) were the least successful: 5/192 (2.6%) passed 5 GCSEs C-A* with EWMS, as did 37/354 (10.4%) ineligible for FSM. The strongest associations with these GCSEs were SEN statements (aOR 0.07, 0.06–0.07), FSM eligibility (aOR 0.39, 0.37–0.41), and epilepsy (aOR 0.60, 0.45–0.80). However, data were unavailable for 15-18% of children, mainly those educated outside mainstream schools, and some co-morbidities. Generalisation of findings to other countries rests with readers.

**Conclusions:** Many children with anomalies from affluent households succeeded. The children left behind lived with poverty <u>and</u> ill-health from congenital anomalies and/or epilepsy. SEN provision mitigated the impact of disadvantage, but poor children with anomalies were unlikely to succeed. †taking maternal Welsh Index of Multiple Deprivation (WIMD) 2014 at birth.

## Introduction

Education outcomes predict life chances [1], achievement [2], quality of life [3], and health [4]. However, ill-health and disability are barriers to educational attainment and optimal school performance [5]. Disability and learning difficulties may emanate from congenital anomalies [6–9]. Major congenital anomalies affected 212.6 per 10,000 (95% confidence intervals [CI] 211.8–213.4) of live or still births in Europe 2005–2023 [10]. The prevalence rates of 20% of major anomalies, including congenital heart disease (CHD) and craniosynostosis, are increasing [11, 12].

Most (97.3%) children with major congenital anomalies survive infancy, due to improved care and surgical interventions: [13–18] 96.9% are alive at 10 years [19]. Accordingly, research is focusing on their life chances. However, overall, their educational attainment is lower than their peers’ [20–30], despite provision for special educational needs (SEN) [24, 31–38].

Children with certain anomalies may also be more likely to experience comorbidities, such as epilepsy, cardiovascular insufficiency [39–42]), cancer and injury [43], and require surgery, hospitalisation and other treatments [44], which may detract from educational attainment [5]. Of the three most common long-term medicated conditions requiring hospitalisation in childhood [45], epilepsy is linked with lower educational attainment [46], but the literature offers no consensus on asthma [47–50] and diabetes [51, 52]. Although educational attainment may be affected by congenital anomalies, intergenerational social and cultural capital may not, suggesting that higher socio-economic status may protect individuals from some adverse consequences of congenital anomalies, and time spent in hospital, away from school [53].

Educational attainment is linked with socio-economic status (SES) [54, 55], and childhood ill-health and disability, which are concentrated in the most deprived communities [56–58]. The proportion of UK children with disability increased from 7.3% in 2004/5 to 11% in 2021/22 [58, 59].

Increasing child poverty [60], the widening attainment gaps at age 16 between those eligible and ineligible for free school meals (FSM) [61], re-organisation of SEN provision [62], and the expanding population of children with anomalies attending mainstream schools necessitate understanding the determinants of educational attainment and targets for support. How biological and socio-economic disadvantages combine and interact, are mitigated by additional support as delivered in routine practice, and whether this generates a profoundly disadvantaged subgroup, is not fully explored. The literature focuses on isolated anomalies [24, 25]. Current evidence is limited by the paucity of large whole-population studies with objective measures of educational attainment, including key covariates, such as socio-economic status (SES) as income, access to services, environment, maternal deprivation, preterm birth [32], hospitalisation, comorbidities, and the impact of SEN provision. We aimed to explore the educational attainment of children with and without major congenital anomalies attending state-funded mainstream schools in Wales at the end of primary and secondary schooling, at ages 11 (KS2) and 16 (KS4) [55,56].

## Methods

This population-based cohort study is reported in accordance with the Reporting of Studies Conducted using Observational Routinely-Collected Data (RECORD) guidelines (see RECORD Statement, other information) [63]. The main data linkage and analytical approaches were published before we accessed data [64, 65].

### Data sources

Routinely collected administrative data were accessed via the Wales national databank, the Secure Anonymised Information Linkage (SAIL) Databank [66, 67]. The maternity, prescribing, hospital admission and socio-economic data of all children born in Wales 01/01/1998–31/12/2007 were selected as described [64, 65]. Children born to mothers resident in Wales were identified from the National Child Community Health Database (NCCHD), which supplied data on maternity and perinatal outcomes. Children’s birth dates, sex, and, where relevant, dates of death, were ascertained from the Annual District Births Extract (ADBE) and the Deaths’ Register (ADDE). Records of eligible infants were linked to the Welsh Demographic Service dataset (WDS) to obtain anonymised information on geographical area of residence and facilitate linkage to the Welsh Index of Multiple Deprivation (WIMD) [68]. Approximately 85% of Welsh primary care practices contribute routinely collected data to SAIL, including all primary care prescriptions, from 86.2% of the population and 86.5% of children, as the Welsh Longitudinal General Practice (WLGP) dataset [69]. To include important medicated comorbidities (epilepsy, diabetes, asthma and cardiac conditions), the study was restricted to this portion of the national population. Data on hospitalisations were obtained from the Patient Episodes Dataset for Wales (PEDW). Table 1 summarises the datasets.

**Table 1.**
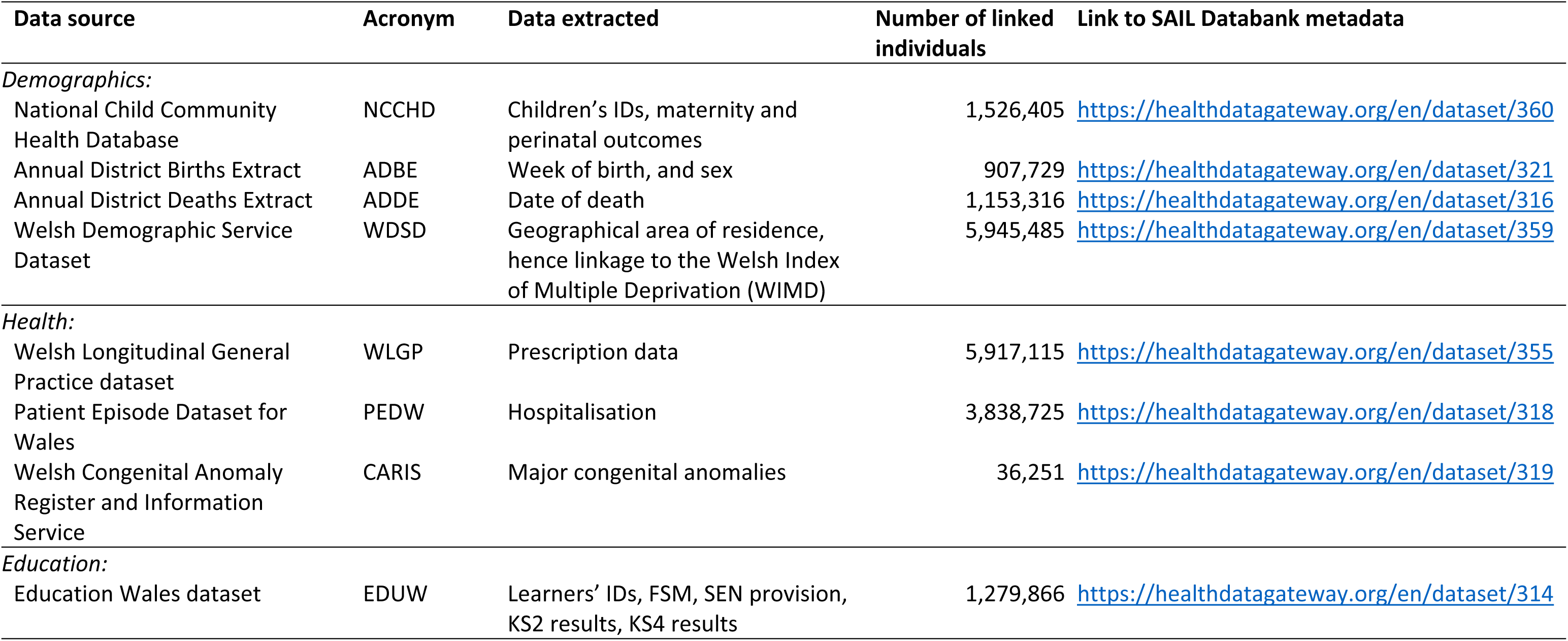
Data in this study.

Major congenital anomalies, as defined and classified by the European Surveillance of Congenital Anomalies (EUROCAT) network [70, 71], are recorded by the Welsh Congenital Anomaly and Information Service (CARIS) register held within Public Health Wales and shared with SAIL. Major congenital anomalies are structural changes that have significant medical, surgical, social or cosmetic consequences for the affected individual, and typically require medical intervention. They account for most of the mortality, morbidity, and disability related to congenital anomalies [71]. Reports of congenital anomalies are received from clinicians, including obstetricians, paediatricians and surgeons, and families, and verified by the database manager (DT). We analysed ‘All anomalies’ as a single group defined by allocation of ICD10 codes listed in EUROCAT guide 1.4 (binary variable al1) [70], in line with experts’ [72] and national reporting [73], to offer an overview of needs, and avoid too many small categories obscuring associations [74]. We reported isolated structural anomalies, often associated with surgical intervention, and two chromosomal anomalies, Turner and syndromes, where numbers permitted (S19 List 1). Isolated anomalies are defined by EUROCAT as a congenital anomaly in one organ system only or with a known sequence where multiple congenital anomalies cascade as a consequence of a single primary anomaly [65]. Children with major anomalies outside these categories were analysed as a single group, ‘other anomalies’, together with children with more than one anomaly.

We linked children to their education records until 31/12/2019 in the Pupil Level Annual School Census and Welsh Examinations Database, held within SAIL as the Education Wales (EDUW) dataset. Within EDUW, children in mainstream state-funded schools are identified through school-derived learner IDs. Data relating to children attending special schools were not available, and therefore our analysis is restricted to children considered sufficiently well, physically and mentally, not to need the expertise available only in special schools. Pupil characteristics including eligibility for free school meals (FSM) and records of SEN provision allocated to the pupil from the beginning of each academic year, along with attainment data for KS2 were obtained from the Pupil Level Annual School Census tables within the EDUW dataset, while attainment data for KS4 were obtained from Welsh Examinations Database tables. Within SAIL, all IDs (including learner IDs) are matched to the standard SAIL anonymised linkage field (ALF) using probability matching based on National Health Service (NHS) number or personal identifiers [66, 67].

### Inclusion criteria

Our cohort comprised all live born children with primary care records, with and without major congenital anomalies born in Wales between 1998–2007 [75], alive on their 16^th^ birthday and with academic results in EDUW by age 11 (KS2) or age 16 (KS4) (PHW 2022). Children alive at 11, but dying before 16 were excluded in all analyses. (In Wales, there are ∼30 deaths each year in children aged 12–17; about 30% are expected deaths [76].) Children born 1998–2007 and 1998–2002 were included in the analysis of KS2 (age 11) and KS4 (age 16) results, respectively.

### Outcomes

The Welsh National Curriculum is unique. It aims to provide a firm foundation in language (English or Welsh/Cymraeg), maths and science (EWMS). At KS2, age 11, the expected level of attainment in Wales is Level 4 or above in all subjects including EWMS, assessed through a mixture of standardised and moderated teacher assessments. At KS4, age 16, the end of compulsory schooling, children work towards externally assessed national qualifications in core (EWMS), foundation and optional subjects. The standard KS4 qualifications are the General Certificates of Secondary Education (GCSEs), although institutions may offer other academic or vocational qualifications, which may or may not be equivalent to whole or part GCSEs. The Welsh Examinations Database records whether a child has achieved Level 1 (G–A*) or Level 2 (C–A*) in at least 5 GCSE subjects or equivalents and whether these included EWMS. Achievement of 5 GCSEs, C–A* with EWMS governs access to some higher education and professional courses, such as nursing.

Expected levels of attainment have remained consistent over time. We selected outcomes at ages 11 and 16, consistent with UK-wide standards [77, 78] (Table 2). The lower-level pass outcome captured all children who met expectations to any extent, whilst the high-level outcome only included children meeting expectations in all three core subjects.

**Table 2.** Attainment Benchmarks.

| Stage | Low-level / other pass / any of: | High- level pass |
| --- | --- | --- |
| Key Stage 2,<br>Age 11 | Level 4 English/ Welsh<br>Level 4 Maths<br>Level 4 Science<br>Level 4 English/Welsh & Maths<br>Level 4 English/Welsh & Science<br>Level 4 Maths & Science<br>Level 4 EWMS | Level 4 EWMS |
| Key Stage 4,<br>Age 16 | Level 1 (G-A* in any 5 GCSEs)<br>Level 2 (C-A* in any 5 GCSEs)<br>Level 1 (G-A* in 5 GCSEs including<br>EWMS)<br>Level 2 (C-A* in 5 GCSEs including<br>EWMS) | Level 2 (C-A* in 5<br>GCSEs, including<br>EWMS) |

*EWMS, English/ Welsh, Maths and Science, GCSE General Certificate of Secondary Education*

### Potential Confounders

We adjusted for covariates related to: the child’s sex and perinatal factors; deprivation indices of both child and mother; hospital stay; long-term conditions (as prescribed medicines); and SEN provision. We categorised gestational age data from the NCCHD as very pre-term (<32 weeks; gestation), pre-term (32 to 36+6 weeks) and term (≥37 weeks) (S1 Table).

We accounted for mothers’ deprivation at time of childbirth at local area level and children’s deprivation at both local area and household level. We took the access to services (ATS), physical environment and income quintile scores from the 2014 version of the WIMD, as measures of the child’s local area deprivation [79], using the child’s registered address on the 1^st^ May in the year of their key stage 2 and 4 assessments. WIMD has 8 components: income, employment, health, education, ‘Access to Services’, community safety, physical environment, housing. Each domain is compiled from a range of indicators. For example, the education outcome data comprises: Key Stage 2 ‘Average Point Score’, Key Stage 4 ‘Capped Point Score’, Key Stage 4 ‘Level 2 Inclusive’, repeat absenteeism, proportion of 18-19-year-olds not entering Higher Education, and proportion of 25-64-year-olds with no qualifications [79]. We used the full WIMD to account for mothers’ deprivation, but only selected components (above) for children’s deprivation. We analysed free school meals (FSM) eligibility (not receipt) during the child’s KS2 & KS4 years, as an indicator of household-level deprivation [80].

Children or young people are considered to have SEN if they have a learning difficulty or disability which requires additional educational provision, over and above routine schooling; this does not always parallel severity of physical disability. Procedures for service allocation are described in the 2002 code of practice [81], which has been revised {Welsh Government, 2021 #253}. Needs are assessed by schools and the local education authority; decisions are sometimes appealed. SEN provision recorded in the year of assessment was taken as a marker for the severity of a child’s difficulties: (1) no provision, (2) school action plan (in-school support), (3) action plan plus (involving external support from, for example, psychologists), (4) statementing (involving additional funded education, health and care plans) [81].

Hospitalisation was measured as the total number of days (with overnight stays) spent in hospital by the 1^st^ May of a child’s KS2 assessment year and between KS2 and 1^st^ May of the KS4 assessment year. Medicated asthma, type 1 diabetes, epilepsy or cardiac conditions, were indicated by primary care records of > 1 prescription for any of these conditions [82] by the 1^st^ May of a child’s key stage 2 and 4 assessment years. ATC (anatomic, therapeutic chemical) codes related to these conditions were identified, as previously [83–85], and matched with Readv3 codes (see ‘Codes’ below). We discounted single prescriptions as likely to represent non-adherence.

### Statistical analyses

Record linkage was undertaken via Structured Query Language. All other data preparation and analyses were completed in R version 4.3.3 using packages: stats v4.3.3, dplyr v1.1.4, olsrr v0.6.0, lmtest v0.9.40.

Congenital anomalies were analysed together, and, where numbers permitted, as isolated anomalies. We tested whether anomalies and FSM altered the likelihood of having SEN provision, and the association between maternal deprivation and anomalies. To explore whether anomalies were associated with educational attainment, when demographic, biological and socio-economic factors were accounted, logistic regression [86] was used to explore four education outcomes (Table 2), set as binary categorical variables. Explanatory variables were selected and categorised *a-priori*. Covariates were added to models in four categories: child traits (year of birth, sex, gestational age), deprivation (FSM, ATS quintile, physical environment quintile, income quintile, mother’s deprivation quintile), medical interventions and co-morbidities (days in hospital, asthma, epilepsy, diabetes, cardiac conditions), and SEN provision (Supplementary file 2). Models were tested for collinearity. To avoid over-specification in these explanatory models (Fig.1), results were reported with and without SEN provision, Down syndrome and deprivation, and separately for boys and girls, and for each of the four levels of SEN provision. We assessed the associations between educational attainment and the combined effects of: a) SEN and anomalies/sex/hospitalisation/deprivation measures in turn and b) anomalies and deprivation measures in turn as interaction terms (Supplementary file 2). A sensitivity analysis was undertaken for FSM eligibility at both key stages: all missing values were imputed as eligible, and then as ineligible [30]. There were no other data imputations.

#### Ethical and governance approval

The EUROlinkCAT study was approved by SAIL’s Independent Governance Review Panel (IGRP), (number 0511), on 27/1/2018 and 20/12/2022. Disclosure of small numbers (1–4) in any category is prohibited, as discussed [87].

## Results

We report on 256,568 children at age 11, KS2, and 128,727 at age 16, KS4 (Table 3). Most (83–87%) children were linked to their education records at both key stages. A further 2–5% children had either no primary care data or no information on SES or uncertain gestation (S1, S2 Tables). After exclusions for missing data, we identified 8,754 children with anomalies with key stage 2 records and 4,427 with key stage 4 records (Table 3, Flow diagram, Fig 2a,b). Most children present in the KS4 cohort were also present in the KS2 cohort: 4,350 children with anomalies and 122,100 children without anomalies contributed to education records at both stages. Children (77 with anomalies and 2200 without) transferring into mainstream education in Wales were present at KS4, but not KS2. Transfers were from private, home or special education. We found no differences in SES between children with and without primary care data (Supplementary information file 1).

**Fig. 2a:**
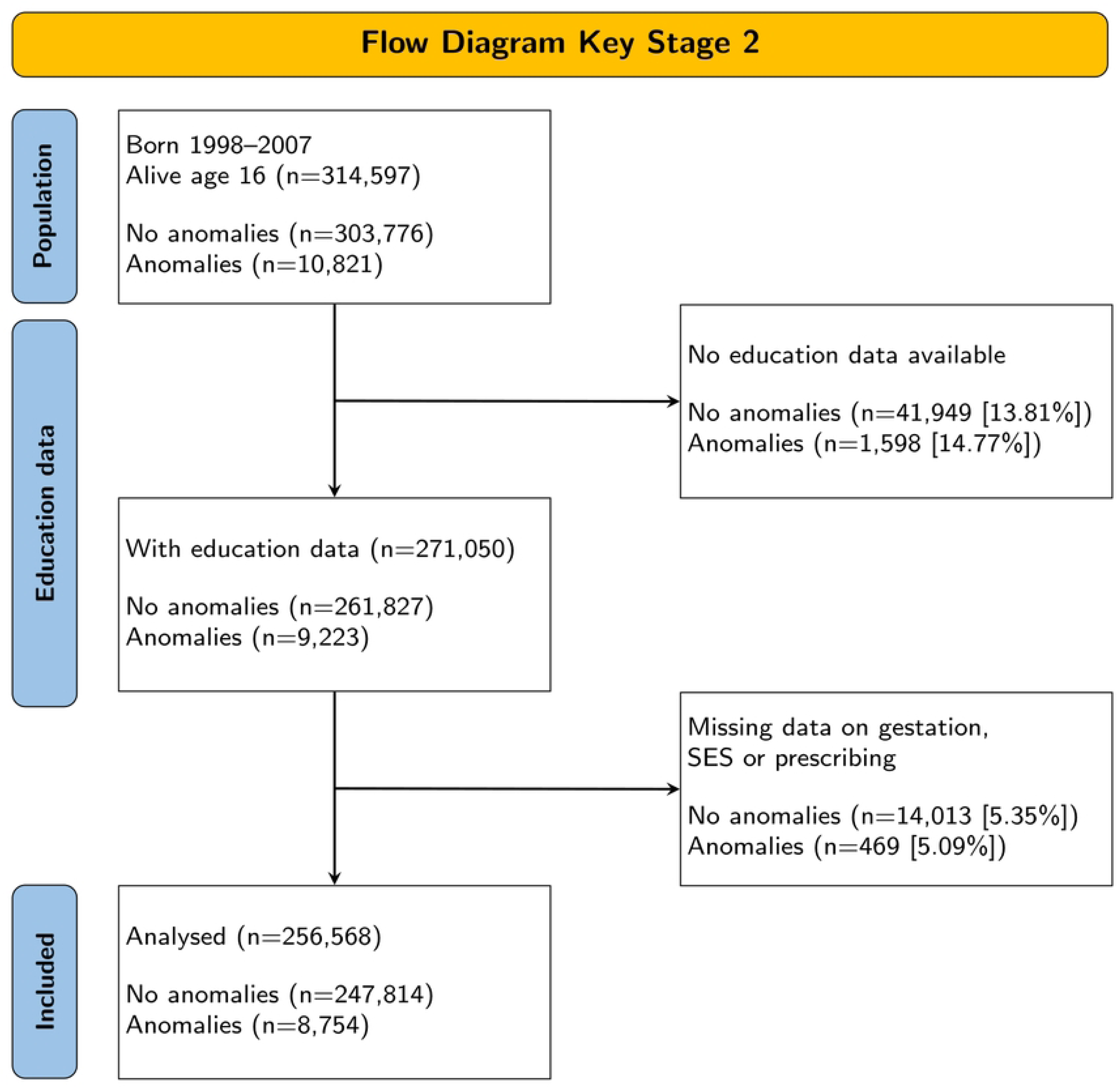
Flow diagram of participants at age 11, key stage 2.

**Fig. 2b.**
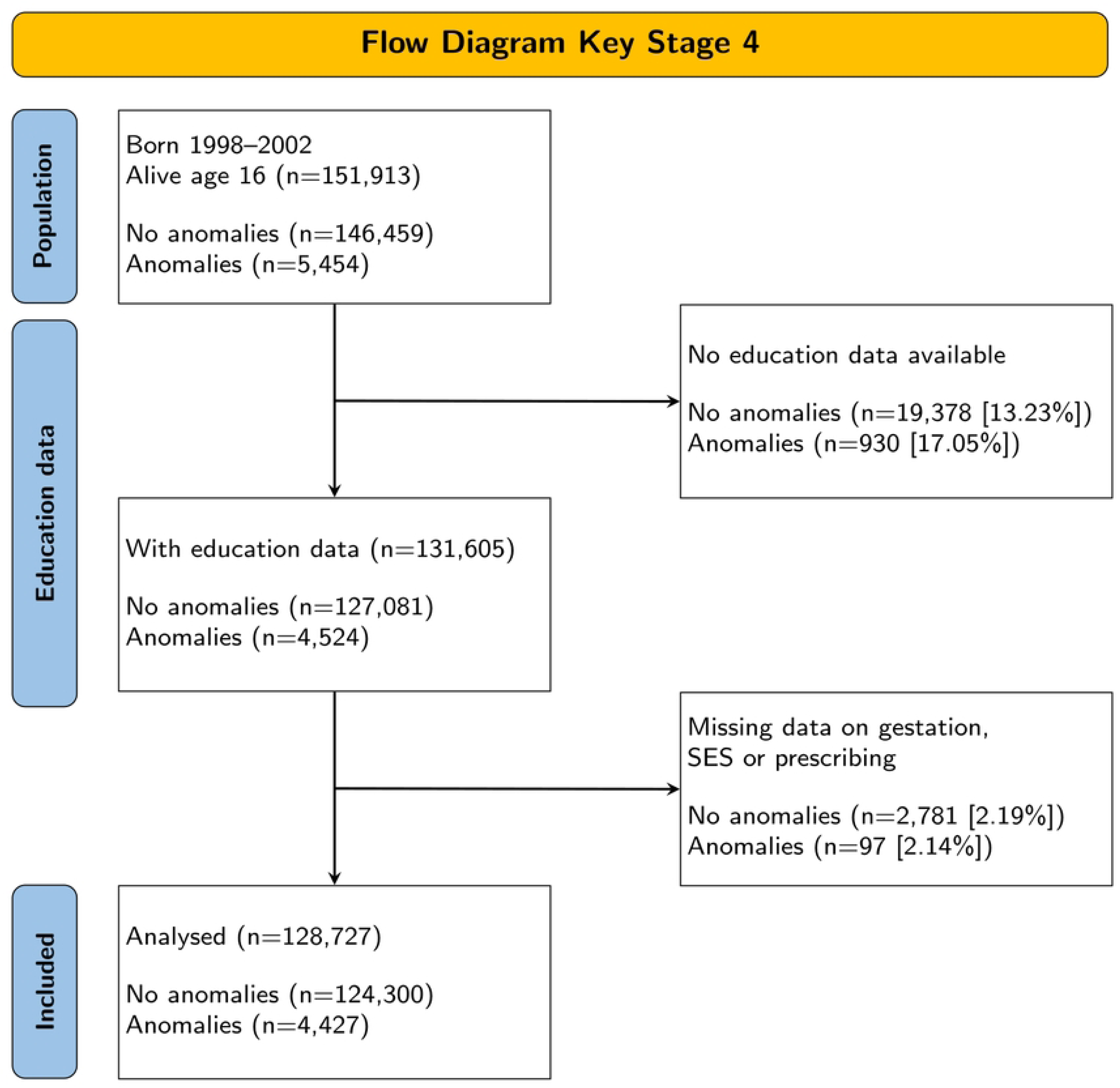
Flow diagram of participants at age 16, key stage 4.

**Table 3.** Numbers in the Cohorts.

|  | Available |  | Linked to attainment data |  |  |  | Linked to all data |  |  |  |
| --- | --- | --- | --- | --- | --- | --- | --- | --- | --- | --- |
|  | With anomalies<sup>§</sup> | No anomalies | With anomalies |  | No anomalies |  | With anomalies |  | No anomalies |  |
| KS2<br>* | 10,821 | 303,776 | 9223 | 85.5% | 261,827 | 87.0% | 8754 | 81.2% | 247,814 | 82.4% |
| KS4<br>† | 5454 | 146,459 | 4524 | 82.9% | 127,081 | 86.8% | 4427 | 81.2% | 124,300 | 84.9% |

Notes to table: *KS key stage*
\* born 1998 – 2007; † born 1998– 2002, %s of total quoted.
<sup>§</sup> children with anomalies included in the al1 category of EUROCAT guide 1.4

Only children identified in all data sets are included in this analysis.

Just under half the children with major anomalies could be categorised within the ‘isolated structural anomalies’ taxonomy. The remainder had more than one anomaly or a genetic or other syndrome or a major anomaly with too few children for analysis (S9 List 1) (Table 4).

**Table 4:** Prevalence of Major Congenital Anomalies.

| Isolated anomalies* | Key Stage 2 (age 11) born 1998-2007 |  | Key Stage 4 (age 16) born 1998-2002 |  |
| --- | --- | --- | --- | --- |
|  | n | % of children with anomalies | n | % of children with anomalies |
| Cleft lip with or without cleft palate | 177 | 2.0% | 78 | 1.8% |
| Cleft palate | 153 | 1.7% | 74 | 1.7% |
| CHD (excluding severe CHD) | 1617 | 18.5% | 808 | 18.3% |
| Severe CHD | 383 | 4.4% | 183 | 4.1% |
| Oesophageal atresia | 14 | 0.2% | <10 | 0.2% |
| Ano-rectal atresia and stenosis | 21 | 0.2% | <15 | 0.3% |
| Diaphragmatic hernia | 26 | 0.3% | 15 | 0.3% |
| Gastroschisis | 119 | 1.4% | 43 | 1.0% |
| Multicystic renal dysplasia | 99 | 1.1% | 48 | 1.1% |
| Hypospadias | 691 | 7.9% | 347 | 7.8% |
| Spina bifida | 22 | 0.3% | 13 | 0.3% |
| Limb reduction defects | 70 | 0.8% | 35 | 0.8% |
| Hydrocephalus | 55 | 0.6% | 28 | 0.6% |
| Craniosynostosis | 99 | 1.1% | 45 | 1.0% |
| Chromosomal anomalies |  |  |  |  |
| Down syndrome† | 202 | 2.3% | 66 | 1.5% |
| Turner syndrome† | 27 | 0.3% | 13 | 0.3% |
| Other EUROCAT anomalies* | 4979 | 56.9% | 2610 | 59.0% |
| All anomalies (EUROCAT al1) ‡ | 8754 | 100% | 4427 | 100% |
|  | % of population |  | % of population |  |
| No anomaly | 247,814 | 96.6 | 124,300 | 96.6 |
| TOTAL | 256,568 | 100 | 128,727 | 100 |

Notes to table. CHD congenital heart defect
\*Anomalies were as defined in EUROCAT guide 1.4 (S19 List 1)
† children with Down or Turner syndrome with additional anomalies are included here, as the problems have a common genetic aetiology.
‡ To delineate congenital anomalies and achieve comparability of prevalence rates between registers across Europe, the count of cases with all anomalies (al1) includes all cases with one or more codes in the Q chapter of ICD10 and a limited set of conditions coded outside the Q chapter, as specified in the definition of subgroups in Chapter 3.3 (EUROCAT 2013/2021 P.81): Q-chapter, D215, D821, D1810^, P350, P351, P371 [70].
Low numbers have been redacted to ensure compliance with governance requirements in subgroup analyses in supplementary tables.

### Cohort description: the burden of poverty

When compared with children without anomalies, children with anomalies were more likely to be male, born pre-term to mothers in the most and least deprived areas, eligible for FSM, living in the most deprived income quintile and most deprived environment quintile, They were less likely to live in the areas with poor service connections. The largest differences in prevalence of deprivation (of ∼2-4%), between those with and without anomalies, related to access to services (ATS) and FSM eligibility. Other prevalence differences were ∼1% (Table 5).

**Table 5.**
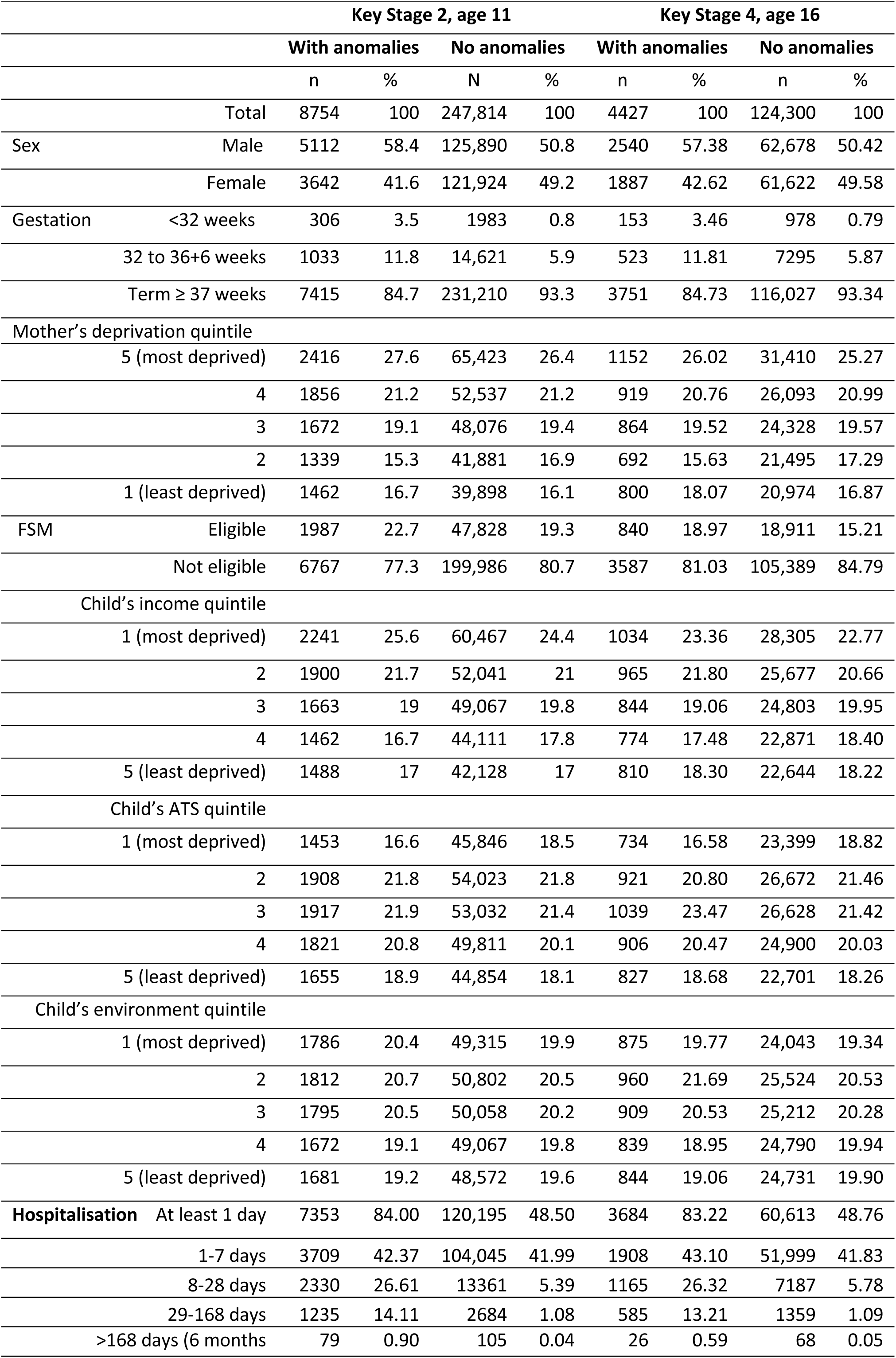
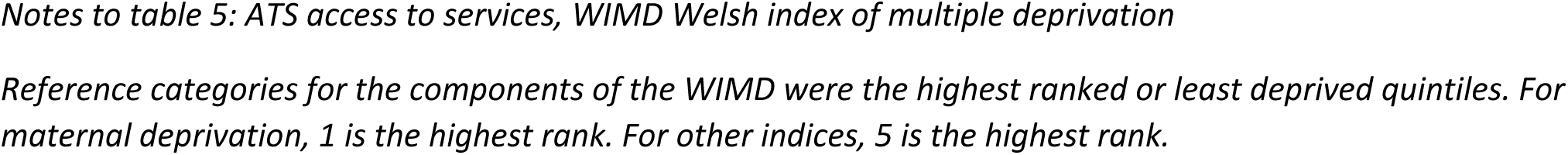
Children with and without congenital anomalies.

|  |  | Key Stage 2, age 11 |  |  |  | Key Stage 4, age 16 |  |  |  |
| --- | --- | --- | --- | --- | --- | --- | --- | --- | --- |
|  |  | With anomalies |  | No anomalies |  | With anomalies |  | No anomalies |  |
|  |  | n | % | N | % | n | % | n | % |
| Total |  | 8754 | 100 | 247,814 | 100 | 4427 | 100 | 124,300 | 100 |
| Sex | Male | 5112 | 58.4 | 125,890 | 50.8 | 2540 | 57.38 | 62,678 | 50.42 |
|  | Female | 3642 | 41.6 | 121,924 | 49.2 | 1887 | 42.62 | 61,622 | 49.58 |
| Gestation | <32 weeks | 306 | 3.5 | 1983 | 0.8 | 153 | 3.46 | 978 | 0.79 |
|  | 32 to 36+6 weeks | 1033 | 11.8 | 14,621 | 5.9 | 523 | 11.81 | 7295 | 5.87 |
|  | Term ≥ 37 weeks | 7415 | 84.7 | 231,210 | 93.3 | 3751 | 84.73 | 116,027 | 93.34 |
| Mother's deprivation quintile |  |  |  |  |  |  |  |  |  |
| 5 (most deprived) |  | 2416 | 27.6 | 65,423 | 26.4 | 1152 | 26.02 | 31,410 | 25.27 |
| 4 |  | 1856 | 21.2 | 52,537 | 21.2 | 919 | 20.76 | 26,093 | 20.99 |
| 3 |  | 1672 | 19.1 | 48,076 | 19.4 | 864 | 19.52 | 24,328 | 19.57 |
| 2 |  | 1339 | 15.3 | 41,881 | 16.9 | 692 | 15.63 | 21,495 | 17.29 |
| 1 (least deprived) |  | 1462 | 16.7 | 39,898 | 16.1 | 800 | 18.07 | 20,974 | 16.87 |
| FSM | Eligible | 1987 | 22.7 | 47,828 | 19.3 | 840 | 18.97 | 18,911 | 15.21 |
|  | Not eligible | 6767 | 77.3 | 199,986 | 80.7 | 3587 | 81.03 | 105,389 | 84.79 |
| Child's income quintile |  |  |  |  |  |  |  |  |  |
| 1 (most deprived) |  | 2241 | 25.6 | 60,467 | 24.4 | 1034 | 23.36 | 28,305 | 22.77 |
| 2 |  | 1900 | 21.7 | 52,041 | 21 | 965 | 21.80 | 25,677 | 20.66 |
| 3 |  | 1663 | 19 | 49,067 | 19.8 | 844 | 19.06 | 24,803 | 19.95 |
| 4 |  | 1462 | 16.7 | 44,111 | 17.8 | 774 | 17.48 | 22,871 | 18.40 |
| 5 (least deprived) |  | 1488 | 17 | 42,128 | 17 | 810 | 18.30 | 22,644 | 18.22 |
| Child's ATS quintile |  |  |  |  |  |  |  |  |  |
| 1 (most deprived) |  | 1453 | 16.6 | 45,846 | 18.5 | 734 | 16.58 | 23,399 | 18.82 |
| 2 |  | 1908 | 21.8 | 54,023 | 21.8 | 921 | 20.80 | 26,672 | 21.46 |
| 3 |  | 1917 | 21.9 | 53,032 | 21.4 | 1039 | 23.47 | 26,628 | 21.42 |
| 4 |  | 1821 | 20.8 | 49,811 | 20.1 | 906 | 20.47 | 24,900 | 20.03 |
| 5 (least deprived) |  | 1655 | 18.9 | 44,854 | 18.1 | 827 | 18.68 | 22,701 | 18.26 |
| Child's environment quintile |  |  |  |  |  |  |  |  |  |
| 1 (most deprived) |  | 1786 | 20.4 | 49,315 | 19.9 | 875 | 19.77 | 24,043 | 19.34 |
| 2 |  | 1812 | 20.7 | 50,802 | 20.5 | 960 | 21.69 | 25,524 | 20.53 |
| 3 |  | 1795 | 20.5 | 50,058 | 20.2 | 909 | 20.53 | 25,212 | 20.28 |
| 4 |  | 1672 | 19.1 | 49,067 | 19.8 | 839 | 18.95 | 24,790 | 19.94 |
| 5 (least deprived) |  | 1681 | 19.2 | 48,572 | 19.6 | 844 | 19.06 | 24,731 | 19.90 |
| Hospitalisation | At least 1 day | 7353 | 84.00 | 120,195 | 48.50 | 3684 | 83.22 | 60,613 | 48.76 |
|  | 1-7 days | 3709 | 42.37 | 104,045 | 41.99 | 1908 | 43.10 | 51,999 | 41.83 |
|  | 8-28 days | 2330 | 26.61 | 13361 | 5.39 | 1165 | 26.32 | 7187 | 5.78 |
|  | 29-168 days | 1235 | 14.11 | 2684 | 1.08 | 585 | 13.21 | 1359 | 1.09 |
|  | >168 days (6 months | 79 | 0.90 | 105 | 0.04 | 26 | 0.59 | 68 | 0.05 |

Notes to table 5: ATS access to services, WIMD Welsh index of multiple deprivation
Reference categories for the components of the WIMD were the highest ranked or least deprived quintiles. For maternal deprivation, 1 is the highest rank. For other indices, 5 is the highest rank.

There was no overall association between maternal WIMD at birth and congenital anomalies (most deprived vs. least deprived quintiles OR 1.03, 0.97–1.10). Only gastroschisis, cleft lip and multicystic renal dysplasia were more prevalent in the most deprived quintile (Table S3). Educational attainment was associated with deprivation in unadjusted analyses and when the child’s sex was accounted (Table S4).

### Congenital anomalies and the burden of illness

Proportionally fewer children were issued prescriptions for asthma, epilepsy, and cardiac conditions at age 16 than at age 11. Children with anomalies were more likely to be hospitalised (and for longer), and receive prescriptions for cardiac conditions, asthma and epilepsy, but not diabetes. Cardiac medicines were largely confined to children with CHD and Down syndrome, and could not be analysed independently. A higher proportion of children with CHD and ‘any anomaly’ were treated for epilepsy. Most children with prolonged hospitalisation did not have anomalies (Table 6).

**Table 6.** Comorbidities amongst children with and without congenital anomalies.

| Anomaly | N | Days hospitalised *<br>Median (25<sup>th</sup>, 75<sup>th</sup> centile) | Asthma-related prescriptions<br>N<br>Prescriptions >1 (%) | Prescriptions for epilepsy<br>N<br>Prescriptions >1 (%) | Insulin prescriptions<br>N<br>Prescriptions >1 (%) | Cardiac-related prescriptions<br>N<br>Prescriptions >1 (%) |
| --- | --- | --- | --- | --- | --- | --- |
| **Key Stage2** |  |  |  |  |  |  |
| ALL ANOMALIES (al1) | 8754 | 5 (1, 16) | 1691 (19.3) | 177 (2.0) | 16 (0.18) | 359 (4.1) |
| Cleft lip ± cleft palate | 177 | 11 (7, 18) | 30 (17.0) | <10 | <10 | 0 |
| Cleft palate | 153 | 7 (5,11) | 29 (19.0) | <10 | 0 | 0 |
| CHD (excluding severe CHD) | 1617 | 3 (1, 11) | 316 (19.5) | 23 (1.4) | <10 | 87 (5.4) |
| Oesophageal atresia | 14 | 40 (17,65) | <10 | 0 | 0 | 0 |
| Ano-rectal atresia and stenosis | 21 | 14 (8, 23) | <10 | 0 | 0 | 0 |
| Diaphragmatic hernia | 26 | 18.5 (10,34) | <10 | 0 | 0 | 0 |
| Gastroschisis | 119 | 33 (22, 47) | 22 (18.5) | 0 | 0 | 0 |
| Multi-cystic renal dysplasia | 99 | 2 (1, 6.5) | 20 (10.2) | <10 | 0 | <10 |
| Hypospadias | 691 | 4 (2, 8) | 126 (18.2) | <10 | 0 | 5 |
| Spina bifida | 22 | 10.5 (1, 33) | <10 | <10 | 0 | 0 |
| Limb reduction defects | 70 | 4 (1, 9.8) | 11 (15.7) | <10 | 0 | 0 |
| Hydrocephalus | 55 | 15 (5, 43) | <10 | <10 | 0 | <10 |
| Craniosynostosis | 99 | 7 (3, 12) | 16 (16.2) | <10 | 0 | 0 |
| Down syndrome | 202 | 13 (4, 33) | 49 (24.3) | <10 | <10 | 30 (14.9) |
| Turner syndrome | 27 | 5 (2, 19) | 0 | <10 | 0 | <10 |
| Severe CHD | 383 | 21 (10, 48) | 76 (19.8) | <10 | 0 | 124 (32.4) |
| Other anomaly | 4979 | 5 (1, 15) | 966 (19.4) | 124 (2.5) | <10 | 110 (2.2) |
| No anomaly | 247,814 | 0 (0, 2) | 39,962 (16.1) | 888 (0.4) | 383 (0.15) | 172 (0.07) |
| **Key Stage4** |  |  |  |  |  |  |
| ALL ANOMALIES (al1) | 4427 | 5 (1, 15) | 608 (13.7) | 43 (1.0) | <10 | 104 (2.4) |
| Cleft lip ± cleft palate | 78 | 10 (7, 19) | 10 (12.8) | 0 | <10 | 0 |
| Cleft palate | 74 | 6 (4, 12) | 12 (16.2) | <10 | 0 | 0 |
| CHD (excluding severe CHD) | 808 | 4 (1, 12) | 112 (13.9) | <10 | <10 | 30 (3.7) |
| Oesophageal atresia | <10 | 39 (15, 56) | <10 | 0 | 0 | 0 |
| Ano-rectal atresia and stenosis | <15 | 12.5 (9, 23) | <10 | 0 | 0 | 0 |
| Diaphragmatic hernia | 15 | 18 (9, 40) | <10 | 0 | 0 | 0 |
| Gastroschisis | 43 | 30 (6, 46) | <10 | 0 | 0 | 0 |
| Multi-cystic renal dysplasia | 48 | 2 (1, 8.3) | <10 | <10 | 0 | 0 |
| Hypospadias | 347 | 4 (2, 9) | 48 (13.8) | <10 | 0 | 0 |
| Spina bifida | 13 | 6 (0, 15) | <10 | 0 | 0 | 0 |
| Limb reduction defects | 35 | 3 (1, 7) | <10 | 0 | 0 | 0 |
| Hydrocephalus | 28 | 15.5 (5, 36) | <10 | <10 | 0 | <10 |
| Craniosynostosis | 45 | 7 (2, 13) | <10 | 0 | 0 | 0 |
| Down syndrome | 66 | 9.5 (3, 24) | 13 (19.7) | <10 | <10 | <10 |
| Turner syndrome | 13 | 5 (3, 15) | 0 | <10 | 0 | 0 |
| Severe CHD | 183 | 21 (9, 44) | 28 (15.3) | <10 | 0 | 37 (20.2) |
| Other anomaly | 2610 | 4 (1, 14) | 350 (13.4) | 28 (1.1) | <10 | 32 (1.2) |
| No anomaly | 124,300 | 0 (0, 2) | 14,586 (11.7) | 301 (0.24) | 148 (0.12) | 42 (0.03) |

Notes to table 6. Lq lower quartile (25<sup>th</sup> centile), Uq upper quartile (75% centile), CHD congenital heart defect
\*Days in hospital by 1<sup>st</sup> May in the year of the key stage assessment.

### Congenital anomalies and educational attainment in mainstream state-funded schools

In all: 93.4% (239,617/256,568) and 86.5% (222,011/256,568) children met academic standards for low- and high-level passes at KS2; 94.3% (121364/128,727) obtained 5 GCSEs; and 55.78% (71,802/128,727) obtained 5 GCSEs including EWMS at grades C-A*.

Anomalies were associated with significantly reduced likelihoods of attainment in unadjusted analyses, particularly in the low-level or ‘any’ pass model for KS2 (OR 0.30, 0.28-0.32). Effects were less marked at KS4, particularly for 5 GCSEs grades C to A* (KS4 OR 0.69, 0.65-0.73). Inclusion of SEN provision in the regression models reduced, but did not eliminate, the associations between anomalies and lower achievement (Table 7).

**Table 7.** Congenital anomalies and attainment.

|  |  | Children with anomalies |  | Children without anomalies |  | Unadjusted | Adjusted as below | Adjusted as below plus SEN provision |
| --- | --- | --- | --- | --- | --- | --- | --- | --- |
|  |  | N | % | N | % | OR (95% CI) | aOR (95% CI) | aOR (95% CI) |
| KS2 | Pass (any) | 7167 | 81.9 | 232,450 | 93.8 | 0.30 (0.28, 0.32) | 0.36 (0.34, 0.39) | 0.72 (0.66, 0.78) |
| Age | no pass | 1587 | 18.1 | 15,364 | 6.2 |  |  |  |
| 11 | total | 8754 |  | 247,814 |  |  |  |  |
|  | Pass (high) | 6413 | 73.3 | 215,598 | 87.0 | 0.41 (0.39, 0.43) | 0.48 (0.45, 0.51) | 0.83 (0.77, 0.89) |
|  | no pass | 2341 | 26.7 | 32,216 | 13.0 |  |  |  |
|  | total | 8754 |  | 247,814 |  |  |  |  |
| KS4 | 5 GCSEs |  |  |  |  |  |  |  |
| Age | Attained | 3900 | 88.1 | 117,464 | 94.5 | 0.43 (0.39, 0.47) | 0.48 (0.44, 0.53) | 0.78 (0.70, 0.87) |
| 16 | Not attained | 527 | 11.9 | 6836 | 5.5 |  |  |  |
|  | total | 4427 |  | 124,300 |  |  |  |  |
|  | 5 GCSEs with EWMS grades C-A* |  |  |  |  |  |  |  |
|  | Attained | 2070 | 46.8 | 69,732 | 56.1 | 0.69 (0.65, 0.73) | 0.73 (0.69, 0.78) | 0.93 (0.87, 1.00) |
|  | Not attained | 2357 | 53.2 | 54,568 | 43.9 |  |  |  |
|  | total | 4427 |  | 124,300 |  |  |  |  |

Notes to Table 7. EWMS, English/ Welsh, maths and science, GCSE, General Certificate of Secondary Education, OR odds ratio, aOR adjusted odds ratio, LCI lower confidence interval, UCL upper confidence interval, using 95% confidence intervals, KS key stage, SEN special educational needs

### Special Education Needs (SEN) Provision

Children with anomalies were more likely to receive SEN provision than their peers, particularly the more intensive interventions (Table 8).

**Table 8.**
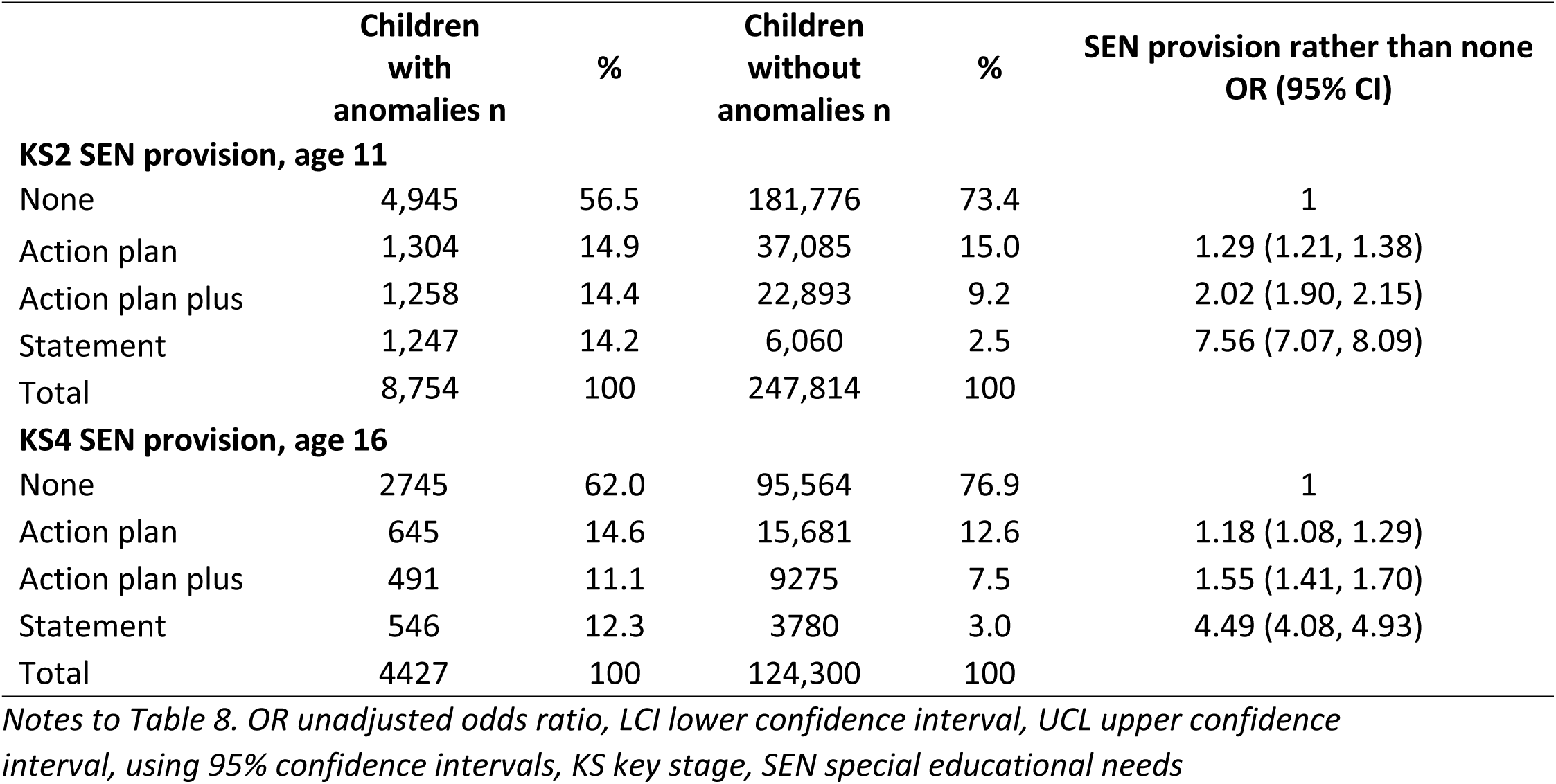
Congenital anomalies and SEN.

Children with anomalies, particularly Down or Turner syndromes, hydrocephalus, *spina bifida*, limb reduction, craniosynostosis, severe CHD (S5a Table), or eligible for FSM were more likely to receive SEN statements (aOR 3.17, 3.00-3.34 at KS2, and aOR 3.97, 3.70-4.26 at KS4) (S5b Table). Adjusting for deprivation made little difference to associations between anomalies and statementing at KS2 (aOR 7.65, 7.15-8.19), and KS4 (aOR 5.06, 4.58-5.60) (S5b Table). However, not all anomalies were statistically significantly associated with statementing (oesophageal atresia, ano-rectal atresia, diaphragmatic hernia, and at KS4 only, gastroschisis, craniosynostosis, multicystic renal dysplasia), but numbers in cross-tabulations were low (S5a table).

Children receiving SEN provision were less successful. Differences in the proportion of children passing were greater between children with /without FSM eligibility than between those with /without congenital anomalies. For 5 GCSEs C-A* with EWMS, for children without SEN support, the pass rate was 30% lower for children eligible for FSM, and 3.6% lower for children with anomalies. For these GCSEs, children with anomalies receiving SEN action plans or ‘action plans plus’ were more successful than their contemporaries receiving the same provision.; SEN provision narrowed the gaps in pass rates between children with/without anomalies and those eligible/ineligible for FSM, only for this outcome, suggesting that it was only a moderator for the highest level of attainment (Table 9).

**Table 9.**
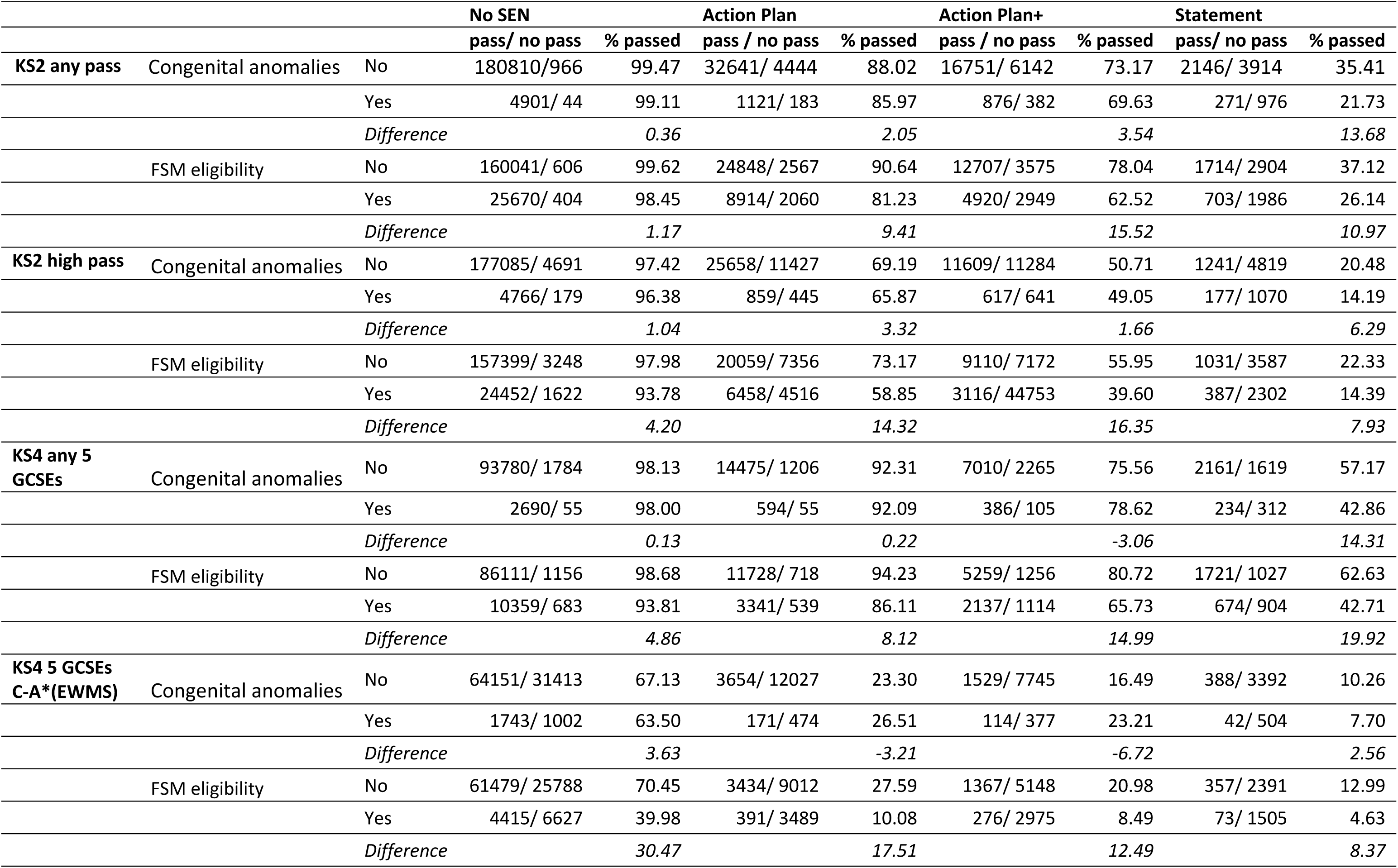
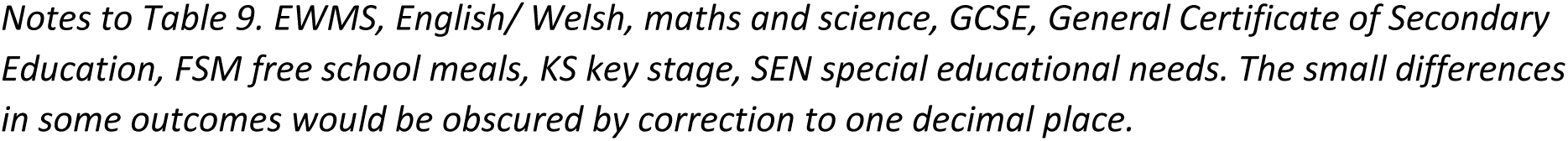
Success and support: congenital anomalies and free school meals eligibility.

|  |  |  | No SEN |  | Action Plan |  | Action Plan+ |  | Statement |  |
| --- | --- | --- | --- | --- | --- | --- | --- | --- | --- | --- |
|  |  |  | pass/ no pass | % passed | pass / no pass | % passed | pass / no pass | % passed | pass/ no pass | % passed |
| KS2 any pass | Congenital anomalies | No | 180810/966 | 99.47 | 32641/ 4444 | 88.02 | 16751/ 6142 | 73.17 | 2146/ 3914 | 35.41 |
|  |  | Yes | 4901/ 44 | 99.11 | 1121/ 183 | 85.97 | 876/ 382 | 69.63 | 271/ 976 | 21.73 |
|  |  | *Difference* |  | 0.36 |  | 2.05 |  | 3.54 |  | 13.68 |
|  | FSM eligibility | No | 160041/ 606 | 99.62 | 24848/ 2567 | 90.64 | 12707/ 3575 | 78.04 | 1714/ 2904 | 37.12 |
|  |  | Yes | 25670/ 404 | 98.45 | 8914/ 2060 | 81.23 | 4920/ 2949 | 62.52 | 703/ 1986 | 26.14 |
|  |  | *Difference* |  | 1.17 |  | 9.41 |  | 15.52 |  | 10.97 |
| KS2 high pass | Congenital anomalies | No | 177085/ 4691 | 97.42 | 25658/ 11427 | 69.19 | 11609/ 11284 | 50.71 | 1241/ 4819 | 20.48 |
|  |  | Yes | 4766/ 179 | 96.38 | 859/ 445 | 65.87 | 617/ 641 | 49.05 | 177/ 1070 | 14.19 |
|  |  | *Difference* |  | 1.04 |  | 3.32 |  | 1.66 |  | 6.29 |
|  | FSM eligibility | No | 157399/ 3248 | 97.98 | 20059/ 7356 | 73.17 | 9110/ 7172 | 55.95 | 1031/ 3587 | 22.33 |
|  |  | Yes | 24452/ 1622 | 93.78 | 6458/ 4516 | 58.85 | 3116/ 44753 | 39.60 | 387/ 2302 | 14.39 |
|  |  | *Difference* |  | 4.20 |  | 14.32 |  | 16.35 |  | 7.93 |
| KS4 any 5 GCSEs | Congenital anomalies | No | 93780/ 1784 | 98.13 | 14475/ 1206 | 92.31 | 7010/ 2265 | 75.56 | 2161/ 1619 | 57.17 |
|  |  | Yes | 2690/ 55 | 98.00 | 594/ 55 | 92.09 | 386/ 105 | 78.62 | 234/ 312 | 42.86 |
|  |  | *Difference* |  | 0.13 |  | 0.22 |  | -3.06 |  | 14.31 |
|  | FSM eligibility | No | 86111/ 1156 | 98.68 | 11728/ 718 | 94.23 | 5259/ 1256 | 80.72 | 1721/ 1027 | 62.63 |
|  |  | Yes | 10359/ 683 | 93.81 | 3341/ 539 | 86.11 | 2137/ 1114 | 65.73 | 674/ 904 | 42.71 |
|  |  | *Difference* |  | 4.86 |  | 8.12 |  | 14.99 |  | 19.92 |
| KS4 5 GCSEs C-A*(EWMS) | Congenital anomalies | No | 64151/ 31413 | 67.13 | 3654/ 12027 | 23.30 | 1529/ 7745 | 16.49 | 388/ 3392 | 10.26 |
|  |  | Yes | 1743/ 1002 | 63.50 | 171/ 474 | 26.51 | 114/ 377 | 23.21 | 42/ 504 | 7.70 |
|  |  | *Difference* |  | 3.63 |  | -3.21 |  | -6.72 |  | 2.56 |
|  | FSM eligibility | No | 61479/ 25788 | 70.45 | 3434/ 9012 | 27.59 | 1367/ 5148 | 20.98 | 357/ 2391 | 12.99 |
|  |  | Yes | 4415/ 6627 | 39.98 | 391/ 3489 | 10.08 | 276/ 2975 | 8.49 | 73/ 1505 | 4.63 |
|  |  | *Difference* |  | 30.47 |  | 17.51 |  | 12.49 |  | 8.37 |

Notes to Table 9. EWMS, English/ Welsh, maths and science, GCSE, General Certificate of Secondary Education, FSM free school meals, KS key stage, SEN special educational needs. The small differences in some outcomes would be obscured by correction to one decimal place.

Of the children with anomalies<u> and</u> FSM eligibility, 2.60% (5/192) receiving SEN statements, and 34.64% (115/332) without SEN provision attained 5 GCSEs C-A* with EWMS. Children with anomalies <u>in</u>eligible for FSM were two to four times more likely to be successful: 10.45% (37/354) of those receiving statements and 67.45% (1628/2413) of those not receiving SEN provision achieved these GCSEs (Fig. 3, S6 Table).

**Fig. 3.**
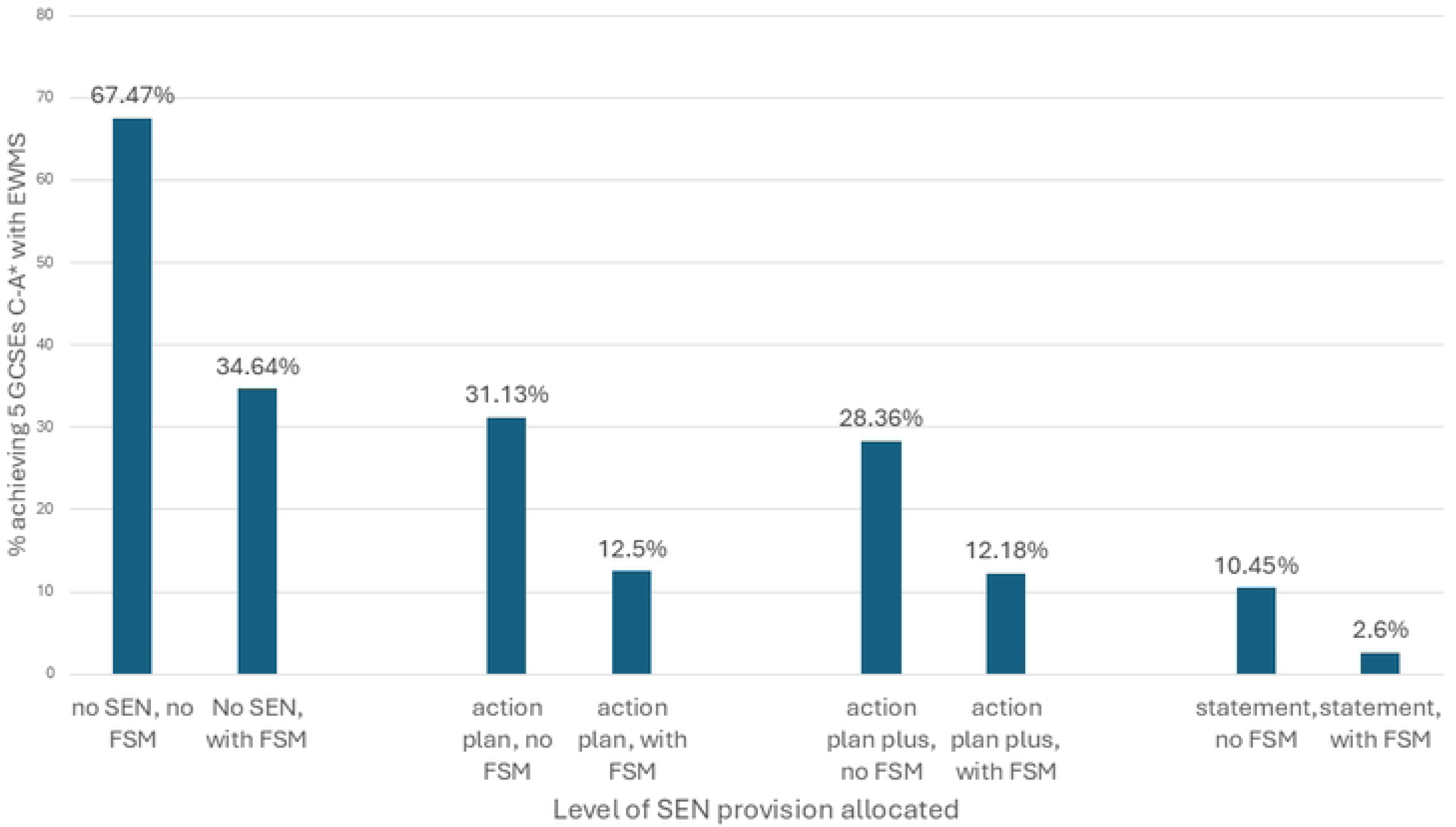
Children with anomalies, with and without FSM eligibility at each level of special needs provision, % achieving 5 GCSEs, C-A* with EWMS.

### Educational attainment: adjusted analyses

Adjusting for preterm birth, sex, deprivation, hospitalisation, co-morbidities (as prescribed medicines), and SEN reduced the association between anomalies and attainment: congenital anomalies were associated with statistically non-significant reduced attainment of 5 GCSEs C-A* with EWMS (aOR 0.93, 0.87-1.003), but associations remained significant for other outcomes (Tables 7,10). Results were similar when children with Down syndrome were removed from the analysis; the association between ‘any anomaly’ and 5 GCSEs C-A* with EWMS was statistically non-significant (aOR 0.94, 0.88-1.01) (S7 table). Removal of SEN provision from the model strengthened the associations between educational attainment and anomalies or epilepsy, but made little difference to the impact of SES (S8 Table). Birth year affected educational attainment, mainly for teacher-assessed KS2 (Table 10). Removing year of birth from the analyses and treating year of birth as a continuous variable did not change the findings; over time, more children were succeeding at age 11, but fewer were attaining GCSEs (Tables S9, S10).

**Table 10.** Associations with attainment at KS2 (n=256,568) and KS4 (n=128,727) (ages 11 and 16)

|  |  | KS2, age 11 |  | KS4, age 16 |  |
| --- | --- | --- | --- | --- | --- |
|  |  | Lower-level pass | Higher level pass | Any 5 GCSEs | 5 GCSEs C-A* with EWMS |
|  |  | aOR (95% CI) | aOR (95% CI) | aOR (95% CI) | aOR (95% CI) |
|  | Any major anomaly | 0.72 (0.66-0.78) | 0.83 (0.77-0.89) | 0.78 (0.70-0.87) | 0.93 (0.87-1.00) |
| Birth year | 1999 | 0.87 (0.81-0.94) | 0.94 (0.88-0.99) | 1.08 (1.00-1.18) | 1.16 (1.12-1.21) |
| Reference | 2000 | 1.03 (0.95-1.11) | 1.21 (1.14-1.28) | 1.09 (1.00-1.19) | 1.18 (1.14-1.23) |
| 1998 | 2001 | 1.28 (1.18-1.38) | 1.56 (1.47-1.66) | 0.90 (0.83-0.98) | 0.98 (0.94-1.02) |
|  | 2002 | 1.39 (1.28-1.50) | 1.95 (1.83-2.07) | 0.84 (0.78-0.92) | 0.93 (0.89-0.97) |
|  | 2003 | 1.58 (1.46-1.72) | 2.42 (2.27-2.57) |  |  |
|  | 2004 | 1.75 (1.61-1.91) | 2.77 (2.60-2.95) |  |  |
|  | 2005 | 1.82 (1.67-1.98) | 3.11 (2.91-3.31) |  |  |
|  | 2006 | 1.89 (1.73-2.05) | 3.36 (3.15-3.58) |  |  |
|  | 2007 | 1.63 (1.49-1.79) | 2.86 (2.66-3.07) |  |  |
| Sex | Male | 1.15 (1.10-1.19) | 1.11 (1.07-1.14) | 0.87 (0.82-0.92) | 0.81 (0.79-0.83) |
| Gestational age | Pre-term 32-36+6 weeks | 1.01 (0.86-1.18) | 0.84 (0.74-0.96) | 1.56 (1.22-2.01) | 0.94 (0.81-1.08) |
| Reference; term | Very pre-term <32 weeks | 0.88 (0.82-0.95) | 0.91 (0.86-0.97) | 1.07 (0.96-1.18) | 1.02 (0.97-1.08) |
|  | FSM eligibility | 0.53 (0.51-0.55) | 0.54 (0.52-0.55) | 0.43 (0.40-0.45) | 0.39 (0.38-0.41) |
| Access to services | ATS1 – most deprived quintile | 1.03 (0.97-1.11) | 0.98 (0.94-1.03) | 0.92 (0.83-1.01) | 0.89 (0.85-0.93) |
|  | ATS2 | 0.91 (0.86-0.97) | 0.94 (0.90-0.98) | 0.90 (0.82-0.98) | 0.86 (0.83-0.90) |
|  | ATS3 | 0.89 (0.84-0.95) | 0.93 (0.89-0.98) | 0.85 (0.78-0.93) | 0.87 (0.84-0.91) |
|  | ATS4 | 0.98 (0.92-1.05) | 0.97 (0.93-1.02) | 0.88 (0.81-0.96) | 0.89 (0.86-0.93) |
|  | Access to services – least deprived quintile | 1 | 1 | 1 | 1 |
| Physical | ENV1 environment – most deprived quintile | 1.12 (1.05-1.18) | 1.02 (0.97-1.06) | 0.92 (0.85-1.01) | 1.03 (0.99-1.07) |
| Environment | ENV2 | 1.13 (1.06-1.20) | 1.06 (1.02-1.11) | 0.94 (0.87-1.03) | 0.98 (0.94-1.02) |
|  | ENV3 | 1.10 (1.03-1.17) | 1.03 (0.99-1.08) | 0.95 (0.87-1.03) | 0.98 (0.94-1.02) |
|  | ENV4 | 1.11 (1.05-1.18) | 1.07 (1.02-1.12) | 1.02 (0.93-1.11) | 1.01 (0.97-1.05) |
|  | Environment 5 – least deprived quintile | 1 | 1 | 1 | 1 |
| Child's household's | INC1 income – most deprived quintile | 0.65 (0.60-0.70) | 0.67 (0.64-0.72) | 0.54 (0.48-0.60) | 0.40 (0.38-0.42) |
| income at | INC2 | 0.70 (0.65-0.76) | 0.75 (0.70-0.79) | 0.61 (0.55-0.69) | 0.48 (0.46-0.50) |
| Assessment | INC3 | 0.80 (0.74-0.87) | 0.81 (0.76-0.86) | 0.74 (0.66-0.83) | 0.59 (0.56-0.62) |
|  | INC4 | 0.94 (0.86-1.02) | 0.90 (0.85-0.96) | 0.86 (0.77-0.97) | 0.74 (0.70-0.77) |
|  | 5 Household income – least deprived quintile | 1 | 1 | 1 | 1 |
| Maternal | 1 – least deprived quintile | 1 | 1 | 1 | 1 |
| Deprivation | 2 – 2<sup>nd</sup> most affluent quintile | 0.99 (0.91-1.08) | 0.98 (0.92-1.04) | 0.90 (0.79-1.01) | 0.85 (0.81-0.89) |
| At birth | 3 – Central quintile for maternal deprivation | 0.85 (0.78-0.92) | 0.85 (0.80-0.90) | 0.82 (0.73-0.92) | 0.72 (0.69-0.76) |
|  | 4 – 2<sup>nd</sup> most deprived quintile | 0.77 (0.71-0.84) | 0.81 (0.76-0.86) | 0.76 (0.67-0.85) | 0.65 (0.62-0.68) |
|  | 5 – most deprived quintile | 0.73 (0.68-0.79) | 0.75 (0.70-0.79) | 0.63 (0.56-0.70) | 0.52 (0.50-0.55) |
| Co-morbidities | Asthma >1 prescription | 1.01 (0.96-1.06) | 1.00 (0.96-1.04) | 1.04 (0.96-1.13) | 0.99 (0.95-1.03) |
|  | Epilepsy >1 prescription | 0.48 (0.41-0.57) | 0.56 (0.47-0.66) | 0.78 (0.58-1.05) | 0.60 (0.45-0.80) |
|  | Insulin >1 prescription | 1.78 (1.18-2.80) | 2.23 (1.59-3.18) | 1.10 (0.59-2.24) | 1.11 (0.76-1.61) |
| SEN | A action plan | 0.05 (0.04-0.05) | 0.06 (0.06-0.07) | 0.29 (0.27-0.31) | 0.17 (0.16-0.18) |
|  | P action plan plus | 0.02 (0.02-0.02) | 0.03 (0.03-0.03) | 0.08 (0.08-0.09) | 0.13 (0.12-0.14) |
|  | S statement | 0.00 (0.00-0.00) | 0.01 (0.01-0.01) | 0.03 (0.03-0.03) | 0.07 (0.06-0.07) |
| Hospitalisation | Each day hospitalised | 1.00 (1.00-1.00) | 1.00 (1.00-1.00) | 1.00 (1.00-1.00) | 1.00 (1.00-1.00) |
| Model | -2LL (log likelihood) | 62,016 | 100,839 | 28,188 | 88,293 |
| Parameters* | Nagelkerke R<sup>2</sup> | 40.2 | 37.9 | 26 | 18.8 |
| | Goodness of fit<sup>†</sup> $\chi^2$ † | 0.4 | 0.38 | 0.26 | 0.19 |

Notes to table 10. ATS access to services, ENV physical environment, EWMS English/ Welsh plus maths and science, FSM free school meals eligibility, GCSE, General Certificate of Secondary Education, KS key stage, INC household income levels, SEN special educational needs, aOR adjusted odds ratio, CI 95% confidence intervals. Odds ratios were adjusted for: sex, gestational age, and year of birth, deprivation, hospitalisation, comorbidities, and SEN provision. Including interaction terms in the models widened confidence intervals, but did not change the findings (S1 file has more details).
Hospitalisation for each day is the only continuous variable in the analysis.
Reference categories for the components of the WIMD 2014 were the least deprived quintile (corresponding with highest rank) [79]. For maternal deprivation, 1 is the least deprived quintile. For other indices, 5 is the least deprived quintile.
- VIF <3 for all models. Income and area-based measures of deprivation were associated, but there was no collinearity between FSM eligibility or gestation age and area-based measures of deprivation. By including all parameters, these models identify the most powerful predictors of educational attainment. There were no data at age 16 for children born 2003-2007.
\*Reported for model comparison
† as described in [88]. Values for $\chi^2$ were statistically non-significant.

For all children, the most marked associations with attainment at KS2 (age 11) were SEN provision, epilepsy, and FSM eligibility; the association with any major congenital anomaly was weaker, particularly at the higher level. At KS4 (age 16), attainment was more strongly associated with FSM than epilepsy or congenital anomalies, particularly when 5 GCSEs C-A* with EWMS were considered. Area-based markers for deprivation, including ‘access to services’, were associated with lower attainment, particularly for 5 GCSEs C-A* with EWMS (Table 10).

For all children educational attainment uniformly followed affluence, leaving the most deprived children with anomalies with little chance of success at age 16/KS4: fewer than 30% of these children attained 5 GCSEs C-A* with EWMS. Children with anomalies living in the least deprived quintiles (fifths) for income (from WIMD) were at least as successful as unaffected children born to mothers in the more deprived quintiles. Children without anomalies from the most deprived quintile were less likely to attain 5 GCSEs C-A* with EWMS than children with anomalies from quintiles 1-3 (Fig. 4) (Table S11).

**Fig 4.**
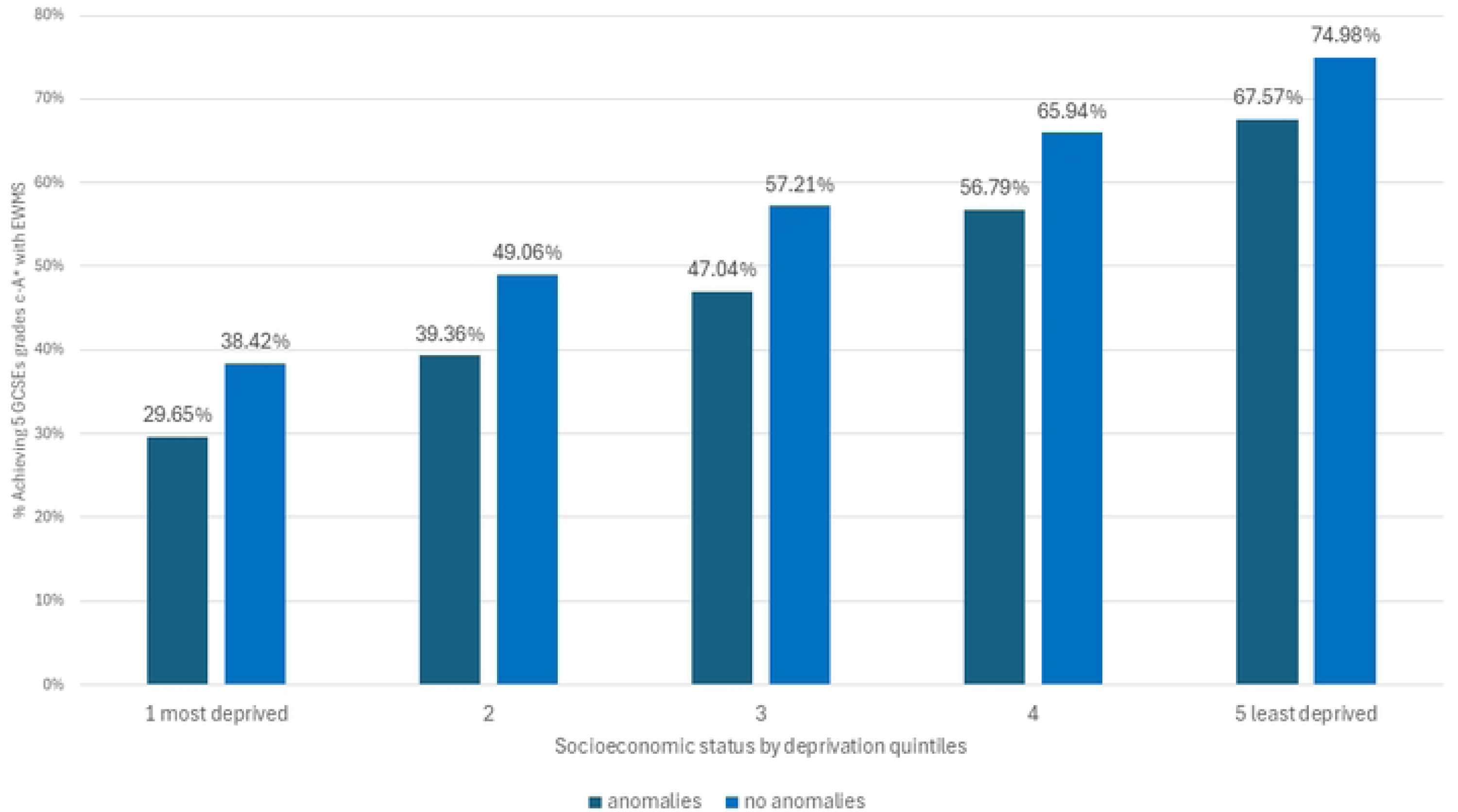
Achieving 5 GCSEs at grades C-A* at age 16, income deprivation and anomalies. Notes to fig.4: Deprivation recorded from the WIMD income domain. WIMD, Welsh Index of Multiple Deprivation.

Although significant at KS2, preterm birth was not associated with attainment at KS4, when SEN was accounted. Hospitalisation (as each day admitted), medicated asthma and type 1 diabetes did not significantly affect outcomes in fully adjusted models. Boys did better than girls at KS2, but only when SEN was accounted. Girls were more successful at KS4 (Tables 10, S8), for some anomalies more than others (Tabes S12a,b). Over time, associations between educational attainment and SEN or FSM changed little at KS4. In unadjusted data, SEN statements were associated with not achieving 5 GCSEs C-A* with EWMS; ORs ranged from 0.047, 0.036, 0.060 for those born in 2002 to 0.061, 0.048, 0.075 for those born in 2001, with intermediate values in other years. At KS2, the negative association between higher pass and SEN statements intensified over time; from 0.012, 0.010-0.015 for children born in 1998 to 0.003, 0.002-0.003 for children born in 2007 (Table S13). Associations between attainment and FSM eligibility intensified over time at KS2: for high pass from 0.325, 0.303-0.348 for birth year 1998 to 0.277, 0.253-0.302 for 2006 and 0.277, 0.250-0.307 for 2007; for low pass from 0.301, 0.275-0.331 for birth year 1998 to 0.242, 0.213-0.275 for 2007 (Table S14).

Inclusion of interaction terms left key findings essentially unchanged; confidence intervals were slightly widened. Most interaction terms including anomalies were not significant, other than the combined effect of SEN school-support with anomalies for KS4 with 5 GCSEs C-A* with EWMS, and statementing combined with anomalies for KS4 any 5 GCSEs. The combined effect of SEN with FSM was significant at all assessments except KS4 with 5 GCSEs C-A* with EWMS; SEN combined with income was significant for some outcomes. In the main, SEN combined with hospitalisation was not significant. Of the WIMD components, only income interacted with SEN, and none interacted with anomalies (S2 file). Findings were not materially altered in sensitivity analysis of the effect of missing FSM eligibility data (S16 Table).

### Isolated congenital anomalies and educational attainment

Children with some, not all, anomalies were vulnerable to failure. In unadjusted analyses, suboptimal educational attainment was associated with Down syndrome, hydrocephalus, CHD, hypospadias or severe CHD, and with gastroschisis and craniosynostosis for most outcomes (S17 Table). After adjusting for SEN provision, FSM eligibility, co-morbidity and hospitalisation, at KS2 (age 11), the effects of CHD, multicystic renal dysplasia, Down syndrome and ‘other anomalies’ remained statistically significant. Down syndrome, gastroschisis and hypospadias were associated with less success in terms of 5 GCSEs at C-A* with EWMS, but associations were lost when boys and girls were analysed separately. The impact of asthma was only apparent at age 16 (KS4). Each day’s hospitalisation had a negative effect at age 11, but this was attenuated by age 16. The impact of FSM eligibility was particularly pronounced on attainment of 5 GCSEs grades C-A*; only Down syndrome and SEN provision were stronger predictors. Insulin use, Turner syndrome, limb reduction, diaphragmatic hernia, *spina biffida,* hydrocepalus, craniosynostosis, and ano-rectal atresia did not adversely affect outcomes at age 16, and *spina biffida had some positive outcomes at age 11,* but numbers with each anomaly in mainstream schools were low. At both stages, epilepsy was more closely associated with poor outcomes than most anomalies (Table 11). Removing FSM from models reduced most associations, particularly with asthma, and one day in hospital was no longer statistically significant (S18 Table).

**Table 11.**
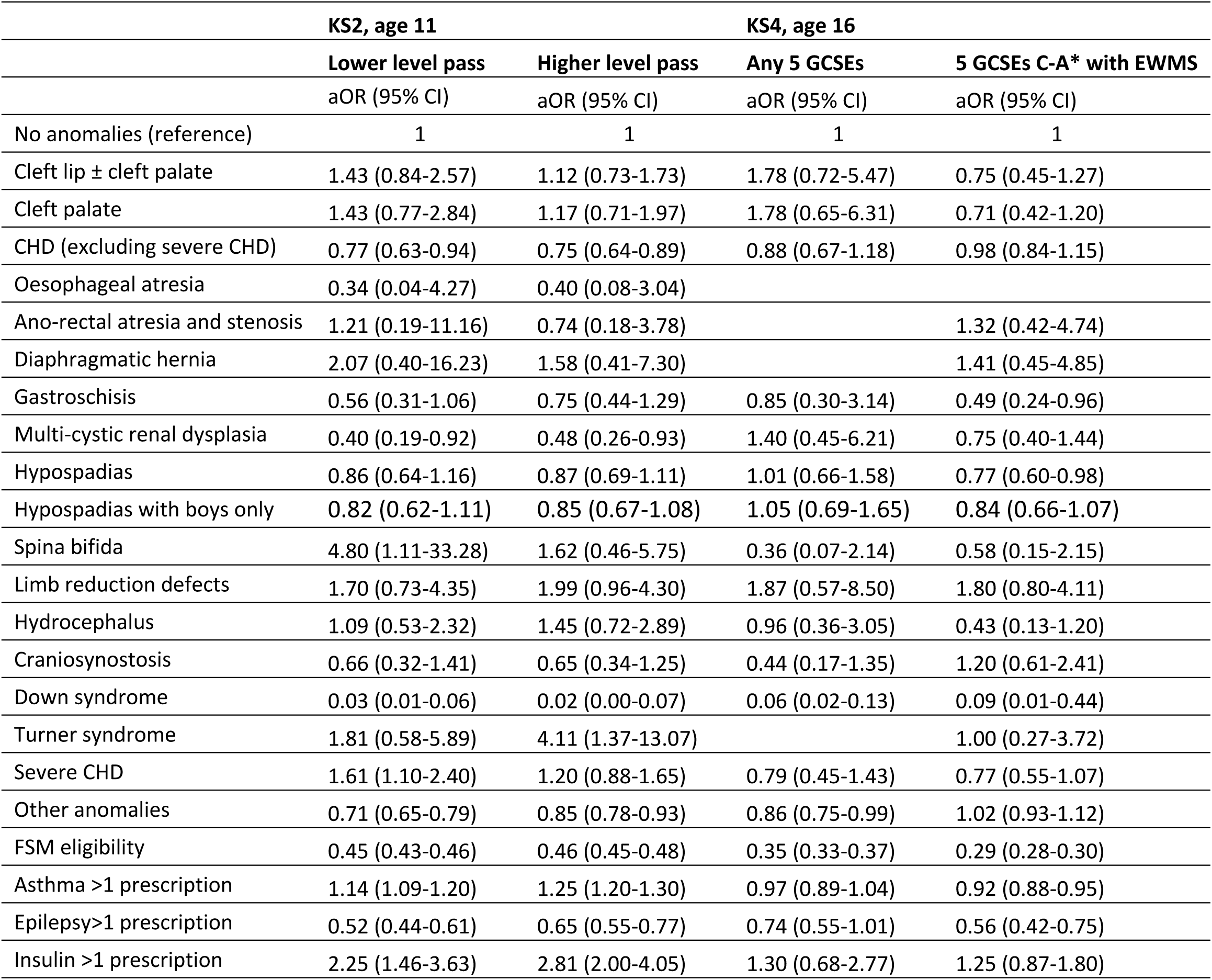
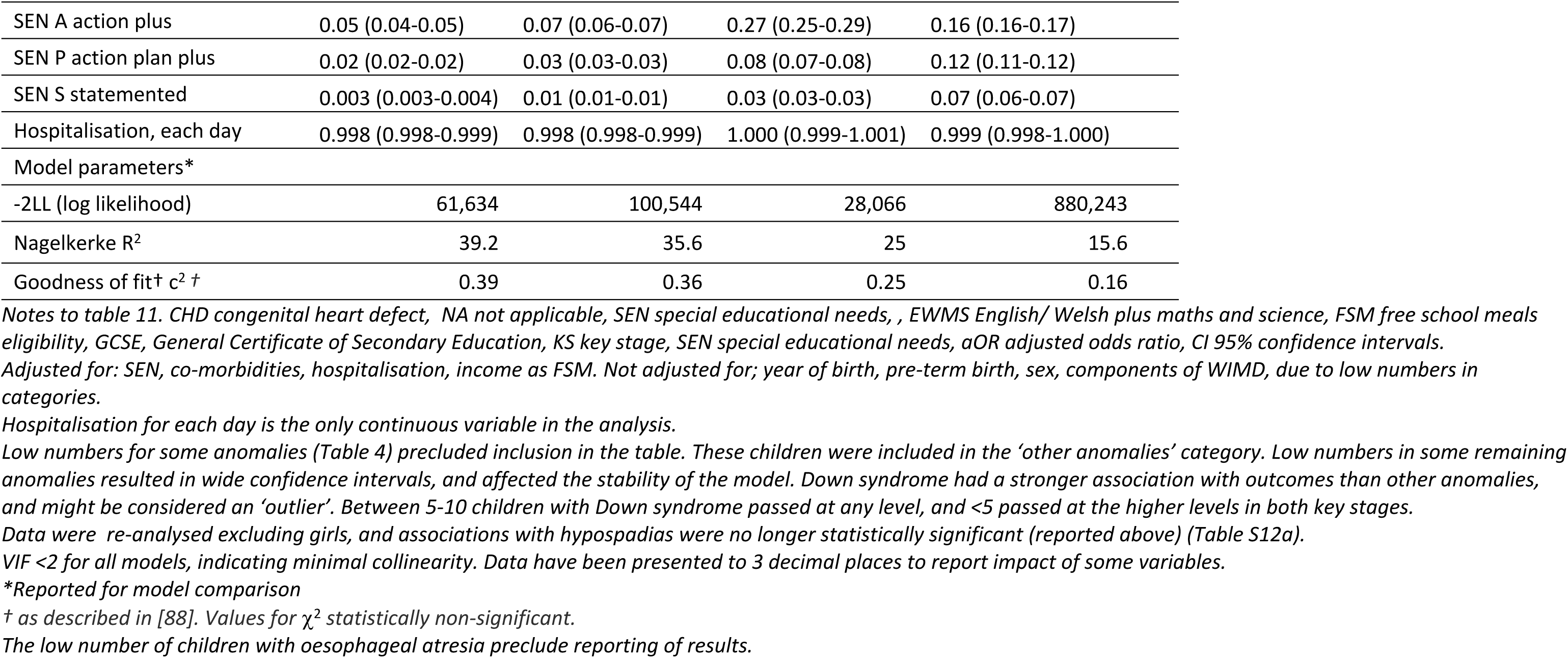
Education outcomes and Congenital anomalies at KS2, age 11 (n=256,568) and KS4, age 16 (n=128,727), adjusted analysis.

|  | KS2, age 11 |  | KS4, age 16 |  |
| --- | --- | --- | --- | --- |
|  | Lower level pass | Higher level pass | Any 5 GCSEs | 5 GCSEs C-A* with EWMS |
|  | aOR (95% CI) | aOR (95% CI) | aOR (95% CI) | aOR (95% CI) |
| No anomalies (reference) | 1 | 1 | 1 | 1 |
| Cleft lip ± cleft palate | 1.43 (0.84-2.57) | 1.12 (0.73-1.73) | 1.78 (0.72-5.47) | 0.75 (0.45-1.27) |
| Cleft palate | 1.43 (0.77-2.84) | 1.17 (0.71-1.97) | 1.78 (0.65-6.31) | 0.71 (0.42-1.20) |
| CHD (excluding severe CHD) | 0.77 (0.63-0.94) | 0.75 (0.64-0.89) | 0.88 (0.67-1.18) | 0.98 (0.84-1.15) |
| Oesophageal atresia | 0.34 (0.04-4.27) | 0.40 (0.08-3.04) |  |  |
| Ano-rectal atresia and stenosis | 1.21 (0.19-11.16) | 0.74 (0.18-3.78) |  | 1.32 (0.42-4.74) |
| Diaphragmatic hernia | 2.07 (0.40-16.23) | 1.58 (0.41-7.30) |  | 1.41 (0.45-4.85) |
| Gastroschisis | 0.56 (0.31-1.06) | 0.75 (0.44-1.29) | 0.85 (0.30-3.14) | 0.49 (0.24-0.96) |
| Multi-cystic renal dysplasia | 0.40 (0.19-0.92) | 0.48 (0.26-0.93) | 1.40 (0.45-6.21) | 0.75 (0.40-1.44) |
| Hypospadias | 0.86 (0.64-1.16) | 0.87 (0.69-1.11) | 1.01 (0.66-1.58) | 0.77 (0.60-0.98) |
| Hypospadias with boys only | 0.82 (0.62-1.11) | 0.85 (0.67-1.08) | 1.05 (0.69-1.65) | 0.84 (0.66-1.07) |
| Spina bifida | 4.80 (1.11-33.28) | 1.62 (0.46-5.75) | 0.36 (0.07-2.14) | 0.58 (0.15-2.15) |
| Limb reduction defects | 1.70 (0.73-4.35) | 1.99 (0.96-4.30) | 1.87 (0.57-8.50) | 1.80 (0.80-4.11) |
| Hydrocephalus | 1.09 (0.53-2.32) | 1.45 (0.72-2.89) | 0.96 (0.36-3.05) | 0.43 (0.13-1.20) |
| Craniosynostosis | 0.66 (0.32-1.41) | 0.65 (0.34-1.25) | 0.44 (0.17-1.35) | 1.20 (0.61-2.41) |
| Down syndrome | 0.03 (0.01-0.06) | 0.02 (0.00-0.07) | 0.06 (0.02-0.13) | 0.09 (0.01-0.44) |
| Turner syndrome | 1.81 (0.58-5.89) | 4.11 (1.37-13.07) |  | 1.00 (0.27-3.72) |
| Severe CHD | 1.61 (1.10-2.40) | 1.20 (0.88-1.65) | 0.79 (0.45-1.43) | 0.77 (0.55-1.07) |
| Other anomalies | 0.71 (0.65-0.79) | 0.85 (0.78-0.93) | 0.86 (0.75-0.99) | 1.02 (0.93-1.12) |
| FSM eligibility | 0.45 (0.43-0.46) | 0.46 (0.45-0.48) | 0.35 (0.33-0.37) | 0.29 (0.28-0.30) |
| Asthma >1 prescription | 1.14 (1.09-1.20) | 1.25 (1.20-1.30) | 0.97 (0.89-1.04) | 0.92 (0.88-0.95) |
| Epilepsy>1 prescription | 0.52 (0.44-0.61) | 0.65 (0.55-0.77) | 0.74 (0.55-1.01) | 0.56 (0.42-0.75) |
| Insulin >1 prescription | 2.25 (1.46-3.63) | 2.81 (2.00-4.05) | 1.30 (0.68-2.77) | 1.25 (0.87-1.80) |
| SEN A action plus | 0.05 (0.04-0.05) | 0.07 (0.06-0.07) | 0.27 (0.25-0.29) | 0.16 (0.16-0.17) |
| SEN P action plan plus | 0.02 (0.02-0.02) | 0.03 (0.03-0.03) | 0.08 (0.07-0.08) | 0.12 (0.11-0.12) |
| SEN S statemented | 0.003 (0.003-0.004) | 0.01 (0.01-0.01) | 0.03 (0.03-0.03) | 0.07 (0.06-0.07) |
| Hospitalisation, each day | 0.998 (0.998-0.999) | 0.998 (0.998-0.999) | 1.000 (0.999-1.001) | 0.999 (0.998-1.000) |
| Model parameters* |  |  |  |  |
| -2LL (log likelihood) | 61,634 | 100,544 | 28,066 | 880,243 |
| Nagelkerke R<sup>2</sup> | 39.2 | 35.6 | 25 | 15.6 |
| Goodness of fit† c<sup>2</sup> † | 0.39 | 0.36 | 0.25 | 0.16 |

Notes to table 11. CHD congenital heart defect, NA not applicable, SEN special educational needs, , EWMS English/ Welsh plus maths and science, FSM free school meals eligibility, GCSE, General Certificate of Secondary Education, KS key stage, SEN special educational needs, aOR adjusted odds ratio, CI 95% confidence intervals.
Adjusted for: SEN, co-morbidities, hospitalisation, income as FSM. Not adjusted for; year of birth, pre-term birth, sex, components of WIMD, due to low numbers in categories.
Hospitalisation for each day is the only continuous variable in the analysis.
Data were re-analysed excluding girls, and associations with hypospadias were no longer statistically significant (reported above) (Table S12a).
VIF <2 for all models, indicating minimal collinearity. Data have been presented to 3 decimal places to report impact of some variables.
\*Reported for model comparison
† as described in [88]. Values for $\chi^2$ statistically non-significant.
The low number of children with oesophageal atresia preclude reporting of results.

At KS4 (age 16), children with diaphragmatic hernia, Down syndrome, hydrocephalus, Turner syndrome, oesophageal atresia, *spina bifida*, limb reduction defects and severe CHD were the most likely to receive SEN statements (S5a Table), but the children least likely to achieve 5 GCSEs at C-A* with EWMS were those with Down syndrome, gastroschisis and hypospadias.

When boys and girls were modelled separately with fewer covariates, only Down syndrome in boys made a statistically significant negative difference at KS4 5 GCSEs at C-A* with EWMS. FSM remained the strongest predictor, followed by epilepsy and asthma. Girls with limb reductions were likely to succeed (Tables S12a,b), contrasting with older reports [21].

In stratified analyses by each category of SEN provision, only when no SEN support was allocated were hospitalisation, asthma, gastroschisis, hypospadias, and (possibly) CHD associated with reduced chances of 5 GCSEs C-A* with EWMS. At all levels of attainment, the FSM association with attainment was stronger amongst children not receiving SEN provision. Epilepsy was more closely associated with attainment amongst children with statements for higher level attainment (S15 Table).

## Discussion

The children least likely to succeed lived in poverty with major congenital anomalies (Fig.3). Educational attainment at age 16 was more strongly associated with deprivation than anomalies or co-morbidities, when hospitalisation, sex, preterm birth, year of birth. and SEN provision were accounted. Accounting for SEN provision only narrowed the attainment gap between children with and without anomalies (Tables 7, 9, S8 Table). The attainment gap between those with and without SEN is narrowing elsewhere [89], but not for children in Wales, whilst the disparity between those eligible and ineligible for FSM is increasing [61, 90].

### Barriers to educational attainment

FSM eligibility, as a measure of individual-level deprivation, was a stronger, if a-theoretical [91], predictor of educational attainment than area-based indices [80, 91]. Children with poor access to services (ATS) were less likely to have anomalies, but less likely to succeed at age 16, highlighting the impact of rurality on educational attainment [92], not reflected in standard deprivation indices [93] and FSM [94, 95].

The category ‘any major congenital anomaly’ encompassed a range of conditions: some anomalies did not statistically significantly detract from performance in adjusted models (table 11). As elsewhere, at various younger ages, children with any major anomaly [30, 96] or CHD [24, 35, 36] had poorer educational attainment than children without anomalies at age 11. Associations between *spina bifida*, craniosynostosis, or diaphragmatic hernia and lower educational attainment reported previously were not confirmed using Wales’ teacher-assessed criteria [20, 22, 24, 26, 28, 31, 33, 34, 97, 98]. Associations with orofacial clefts [14, 20, 22, 26, 27] were confirmed in unadjusted, but not adjusted data (Table 11, Table S17).

At age 16, observations for children with *spina bifida,* diaphragmatic hernia, and CHD were consistent with earlier reports [24, 99–102], with the *caveat* that, whilst many children with *spina bifida* do not complete mainstream schooling, those that do are as successful as their contemporaries [97]. Findings regarding limb reduction defects, hydrocephalus, oesophageal atresia, and Turner syndrome resonate with earlier work [96, 103, 104], whilst findings on anorectal atresia and stenosis, and multicystic renal dysplasia contrast [96, 105, 106]. Disparities in attainment narrowed for most children with anomalies by the end of secondary schooling, age 16 [107, 108], as for other conditions in Wales [109]. However, disparities across the socio-economic spectrum increased, as in children with CHD [110].

Epilepsy was associated with lower attainment, as in Scotland [46]. Adjusted models (including SEN provision) found no impact of prescriptions for asthma [47–50] or diabetes [51, 52] on educational attainment. However, asthma affected attainment at age 16, particularly for children without SEN provision, when some deprivation parameters, including environmental conditions, were not accounted (Table 11). We are unable to separate the impact of illness, including seizures and any underlying brain injury, from effects of school absence or medicines, whether sedative or stimulant.

### Closing the Attainment Gaps

The cumulative impact of deprivation and anomalies left children little chance of achieving the 5 GCSEs needed to access many higher education courses, despite SEN support (Figs 3, 4). Childhood ill-health compounded the effects of poverty; children with anomalies spent more time in hospital [44], and were more likely to be prescribed medicines for epilepsy, asthma and heart conditions (Tables 5,6). Accounting for both SEN and deprivation removed the statistical significance of the association between time in hospital and educational attainment reported elsewhere [34, 111]: this association was only observed for 5 GCSEs C-A* with EWMS amongst children without SEN provision. The detrimental effects of hospitalisation [106] may be mitigated by affluence, where social and cultural capital are available to support learning [55, 112].

Maternal ill-health, obesity, stress, alcohol or substance misuse are all associated with both deprivation [113] and congenital anomalies [114, 115], intensifying disadvantage. Long-term childhood poverty alone depletes social capital, with profound inter-generational consequences. Poverty and low socio-economic status are associated with stress, inadequate stimulation and poor nutrition, all of which adversely affect brain development [116, 117]: abnormalities in frontal and temporal lobes and the hippocampus may account for 15-20% of educational attainment deficits between ages 4 and 22 [118]. Deficits may be exacerbated by: hypoxia induced by cardiac or respiratory anomalies or surgical anaesthesia [7, 119]; anxiety and depression [8]; or autism [9], particularly for children with CHD [8, 120] or receiving corticosteroid therapy [121]. Autism and intellectual delay are associated with CHD [122], long-term conditions [123] or metabolic problems secondary to parenteral nutrition (for gastro-intestinal anomalies) or formula feeding (for children with cleft palate or prolonged hospitalisation) [124, 125]. The absence of overall association between WIMD at birth and congenital anomalies runs counter to reports elsewhere [126–130]; this may be attributed to associations between maternal age and SES in Wales [131]. Biological and socioeconomic disadvantage interact, and are mitigated, but not nullified, by SEN provisions as delivered in routine practice, particularly for children with gastroschisis (Table 11).

Children with the least favourable educational outcomes were not always the most likely to receive the highest level of in-school support (S5a, S5b Tables). For some visible anomalies, such as Turner syndrome [103] or limb reduction defects, additional support may have been instrumental in raising achievement to the level of contemporaries by age 16. We do not know whether the same level of provision might be equally effective for children with anomalies that are less visible after surgery, such as gastroschisis or hypospadias. FSM eligibility did not attract the same investment in SEN as congenital anomalies, despite being more strongly associated with non-attainment of 5 GCSEs C–A* with EWMS (Tables 9, 10,11, S5b Table).

### Strengths and Limitations

To our knowledge, this is one of the largest population cohorts examining academic performance, whilst accounting for holistic life experience: educational support, anomalies, preterm birth, sex, long-term health (epilepsy, diabetes, asthma), hospitalisation, poverty at individual and area level for mother and child, access to services, and the environment. The focus on practical outcomes and analyses of a composite variable of ‘all EUROCAT major anomalies’ [70, 71] minimises uncertainty or obfuscation over coding and classification of disability and perceptions of disability [132]. Analyses of individual anomalies suggests insights into contrasts between visible and less visible anomalies, but low numbers risk obscuring associations [74]. We used individual components of the WIMD score: WIMD has elements relating to education at KS2 & KS4, whose inclusion would have introduced a circular argument [68, 133]. Whole-population data ensures absence of volunteer, and, therefore, collider bias.

Despite the strength of associations between educational attainment, poverty and anomalies, generalisation to unobserved populations or to post-pandemic years, is predicated on logical, not statistical, inference [134, 135]. Education assessments were disrupted during the COVID-19 pandemic, and, currently, there are no more recent education data available. The KS2 teacher assessment has been reviewed, but there are no reasons to question their internal validity [136]; publication of KS2 results ceased in 2018 [137].

This paper describes one European country, where child poverty rates, at 27–36%, are higher than elsewhere in the UK [60] and most OECD countries [138]. However, funding for schools in deprived areas is proportionately lower than in England [139], and expenditure varies between £1,000 and £2,000 per child *per annum* between the 22 local education authorities [140]. The reported prevalence of congenital anomalies is higher than elsewhere in the UK: for example in 2021, England reported congenital anomalies in 235 per 10,000 (231–239), and Wessex in 246 per 10,000 (229–265) of all births [126], whereas Wales reported 319 per 10,000 (299–341) [141]. We do not know whether this discrepancy is due to better case finding, worse air quality [142] in more deprived communities [143], affecting the prevalence of congenital anomalies; [144, 145], poverty [60, 138] or greater use of medicines in pregnancy; [146].

Over 80% of children were linked to their education records. The Pupil Level Annual School Census does not hold data on most independent and special schools; therefore, many children educated privately, at home, outside ‘mainstream’ schools or outside Wales were lost to the study. During primary school, some 2% of children are educated privately and 4.3% in special schools [111]. Incomplete data on special schools disproportionately excludes children with severe disabilities. Provision for some children needing specialist input (for example for blindness or profound deafness) is in England, and data are lost to Wales. However, the anomalies included represent a range of disability, and indicated conditions that might be targeted for additional support (gastroschisis, hypospadias). The omission of prescribing data reduced the size of the cohort, but not its representativeness (file S1). Missing data were largely confined to the outcome variable and missing data for other variables were <5% [147]. Since imputation may underestimate variability whilst never fully compensating for missing data in cohort studies [148], no imputations were undertaken. This complete case analysis is not transferable to children outside mainstream education.

Data on school attendance, social care interventions [149], ethnicity, breastfeeding [150], multiple births, autism, ADHD, mental health, cystic fibrosis, maternal ill-health, medication use or substance misuse [151] were not included [65]. The SAIL Databank does not hold information on fathers, behaviour disturbance or perceptions of bullying, stress or isolation. Primary care records do not document medication adherence; where repeat prescriptions are issued, non-adherence is less likely [152]. To mitigate the difficulties of identifying all illness in electronic health records, and take some account of school attendance, we accounted for time spent in hospital. School attendance may be a mediating covariate [102], and time in hospital accounts for a small proportion of absence. We adjusted for the three most common long-term childhood physical health problems [45, 153]. Rarer conditions were unaccounted, including cerebral palsy or injuries. By using proxies for unmeasured confounders, such as attendance and ill-health outside the three named conditions, we risked overadjustment by controlling for intermediate variables [154]. Our explanatory analyses aimed to identify modifiable risk factors, where interventions might be targeted; propensity score matching might have obscured risk factors or left variables excluded from the propensity score unbalanced, confounding effect estimates [155]. EUROCAT does not assess severity of lesions, but this may not be the only determinant of need [122].

EUROCAT coding gives ‘cleft lip +/- cleft palate’ and ‘cleft palate’, with no ‘cleft lip only’ category. We are, therefore, unable to report on isolated cleft lip, a condition for which there is uncertainty regarding school performance [24]. EUROCAT excludes minor anomalies; however, most do not cause long-term disability, and their impact may be less pervasive.

### Implications for children with health, social <u>and</u> educational needs

Achieving the goals of ‘no child left behind [156], no community left behind’ will entail identifying and ***triaging*** children at risk from compounded socio-economic <u>and</u> biological disadvantage, ensuring they are not overlooked in service re-organisation [62], and expediting intervention [157, 158]. Differences between rich and poor are unsurprising [139]: it is the two- to four-fold magnitude of the differences that should stimulate change. Allocated support, whilst beneficial, is insufficient for children with anomalies from deprived backgrounds to match their contemporaries and for poor children without anomalies to match children with anomalies from relatively affluent families. Routes to educational attainment are needed to counter the increasing impact of poverty [159, 160], low SES [157], and disability, but only intensive programmes will succeed [161, 162].

Children with physical ill-health need holistic specialist care to minimise pain, vision impairment, seizures or hypoxic episodes and compensate for learning difficulties, time spent in hospital [123, 163, 164], and the stress of living in poverty with poor access to services (ATS) [118]. However, the help needed [37], is not always available or accessible due to time and access (ATS) constraints [165]. Would transcending multi-agency provision from education, social and health services, with a unified, holistic, equitable, expert service delivered in specialist on-site units, support the most vulnerable children through school to employment [46, 151, 163, 166–168], and pre-empt poverty, isolation and disability generating a profoundly disadvantaged subgroup?

## Data Availability

All data related to these analyses are contained within the paper and supplementary information. This study uses anonymised data held in the Secure Anonymised Information Linkage (SAIL) Databank. The SAIL Databank independent Information Governance Review Panel (IGRP) approved the study as part of SAIL project 0511. The SAIL Databank trusted research environment can be accessed following a successful application at: https://saildatabank.com.

## Abbreviations

aOR: Adjusted odds ratio
ATS: Access to services
CHD: Congenital heart defect
CI: Confidence intervals / limits
EDUW: Education data for Wales
EWMS: English or Welsh/ Cymraeg (as first language), maths and science
FSM: Free school meals
GCSE: General Certificate of Secondary Education
KS: Key stage
OR: Odds ratio
SAIL: Secure Anonymised Information Linkage
SEN: Special educational needs
SES: Socio-economic status
WIMD: Welsh Index of Multiple Deprivation

## Codes used in this study

Congenital Anomalies were coded according to EUROCAT’s ICD10 codeing published as cited: EUROCAT. Chapter 3.3: EUROCAT Subgroups of Congenital Anomalies (version 23.09.2016). EUROCAT Guide 14: https://eu-rd-platform.jrc.ec.europa.eu/system/files/public/JRC-EUROCAT-Full%20Guide%201%204%20version%2022-Nov-2021.pdf

Coding of medicines used in this study is available in the public domain, in the phenotype library: https://conceptlibrary.saildatabank.com/phenotypes/PH1909/version/3996/detail/

The databases below are unique to Wales and confidential. Data extraction algorithms can be sent on request.

Deprivation: WIMD – we followed the WIMD 2014 guidelines [79].

Education & SEN variables were taken from the education database, as described in the EDUW dataset metadata: https://healthdatagateway.org/en/dataset/314

Births were as recorded in the Annual District Birth Extracts (ADBE), which are based on the national birth register: https://healthdatagateway.org/en/dataset/321

Related parameters were as recorded in the National Community Child Health Dataset (NCCHD): https://healthdatagateway.org/en/dataset/360

## Supporting information

S1 Appendix: Additional details of methods

S1 Table. Gestational age categories

S2 Table Missing data

S3 Table Maternal deprivation and congenital anomalies

S4 Table Education results and deprivation

S5a Table. Associations between isolated anomalies and SEN provision at statement level (unadjusted analysis)

S5b Table. Associations between isolated anomalies and SEN provision at statement level, adjusted for deprivation

S6 Table. Data underlying figure 3. Achieving 5 GCSEs at grades C-A*, children with anomalies, with and without FSM

S7 Table Re-analysis with Down syndrome excluded

S8 Table. Associations with attainment at ages 11 and 16, modelled without SEN provision

S9 Table Re-analysis without year of birth

S10 Table Re-analysis with year of birth as a continuous variable

S11 Table. Data underlying figure 4: achieving 5 GCSEs at grades C-A* with EWMS at age 16, maternal deprivation and anomalies

S12a Table Re-analysis including boys only

S12b Table Re-analysis including girls only

S13 Table Impact of SEN birth years 1998-2007

S14 Table Impact of deprivation birth years 1998-2007

S15 Table education attainment stratified by SEN provision

S16 Table sensitivity analysis, for deprivation data

S17 Table. Individual congenital anomalies and educational attainment, unadjusted data.

S18 Table. Education outcomes and anomalies, adjusted for comorbidities and SEN, modelled without deprivation

S19 List 1. EUROCAT anomalies in this study as in EUROCAT Guide 1.4 S2 Appendix Supplementary file: Analyses including interaction terms

**Other information: RECORD Statement**

---

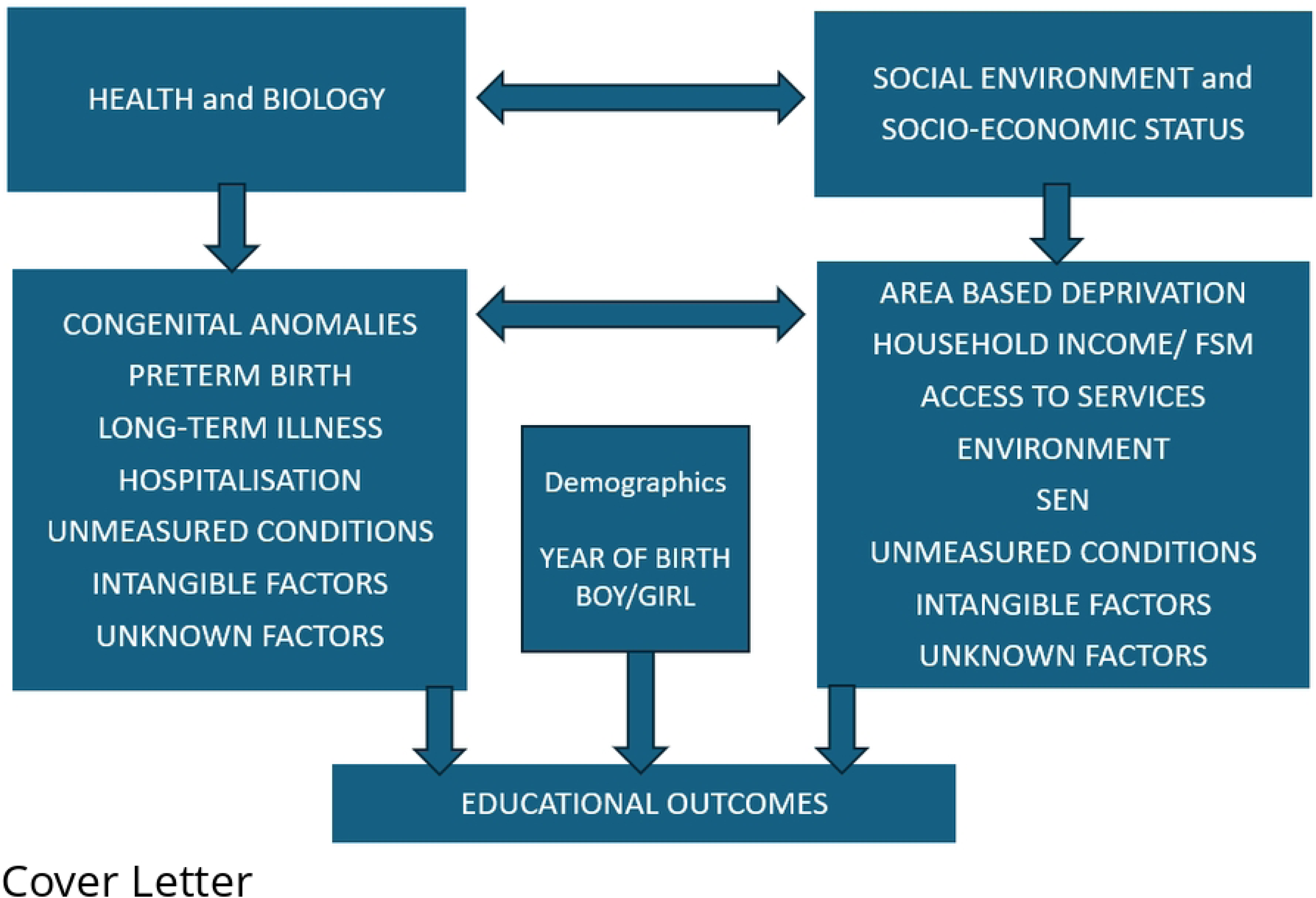

